# Effects of Omega-3 Fatty Acid Treatment on Risk for Atrial Fibrillation: An Updated Meta-Analysis of 34 Trials including 114,326 Individuals

**DOI:** 10.64898/2025.12.14.25342167

**Authors:** Nada R. Abuknesha, James H. O’Keefe, Frank Qian, Nathan L. Tintle, Yidie Lin, Yue Sun, Han-Zhu Qian, Paul S. Aisen, Christine M. Albert, William J. Aronson, Abdul Aziz A. Asbeutah, Heike A. Bischoff-Ferrari, Matthew J. Budoff, Nicholas R. Burns, Cecilia A. Cardenas, Cynthia M. Carlsson, Emily Y. Chew, Neal J. Cohen, Léopold K. Fezeu, Amana Liddell, Pilar Galan, Mark A. Hull, Tsuo-Hung Lan, Pan-Yen Lin, Alexia Mengelberg, Anne-Marie Minihane, Joseph F. Quinn, Thomas A.B. Sanders, David A. Schoenfeld, Andrew Scholey, Kirsty Sprange, Kuan-Pin Su, Christopher H. van Dyck, Carol A. Van Hulle, David Vauzour, Claire Weber, Francine K. Welty, Gary Wittert, Salim Yusuf, William S. Harris

## Abstract

**Background:** Recent meta-analyses of randomized controlled trials (RCTs) have raised concerns that treatment with omega-3 fatty acids may increase risk of atrial fibrillation (AF). However, these meta-analyses included at most eight trials. The aim of this current meta-analysis was to expand the search by including other eligible omega-3 RCTs with AF incidence data, incorporating both published and unpublished data.

**Methods:** Eligible studies were RCTs investigating daily doses of ≥500 mg/d of docosahexaenoic acid (DHA) and/or eicosapentaenoic acid (EPA). Additional inclusion criteria included ≥12 months treatment with EPA/DHA, participants ≥ 50 years of age, and where possible, the absence of known AF/atrial flutter at baseline. The primary outcome was occurrence of new-onset AF. Our primary hypothesis was that risk for AF would simultaneously depend on both omega-3 dose (above or below 1500 mg/d) and background cardiovascular disease (CVD) risk status, and that their combined impact on AF risk would be synergistic.

**Results:** A total of 34 RCTs (36 datasets; n=114,326) were included in this meta-analysis. Only studies including patients at high-risk for CVD who were treated with high-doses of EPA/DHA (>1500 mg/day) showed a statistically significant increase in AF risk with a pooled odds ratio (OR) of 1.48 (95% CI, 1.21-1.81) and an absolute risk difference of 0.8% (0.40-1.1%). None of the other three groups showed statistically significant levels of AF risk (ORs 1.07 (high risk, low dose), 1.06 (low risk, low dose) and 0.95 (low risk, high dose).

**Conclusion:** This meta-analysis suggests that treatment with EPA/DHA is most likely to increase risk for AF in patients at high-risk for CVD who are treated with high doses of EPA/DHA. The risk for AF should be balanced against the benefits of EPA/DHA in making treatment decisions.

## Introduction

Recent meta-analyses of randomized controlled trials (RCTs) have raised concerns that omega-3 fatty acid (FA) supplementation may increase the risk of incident atrial fibrillation (AF), potentially in a dose-dependent manner. Gencer et al. reviewed seven RCTs focused on cardiovascular (CV) outcomes and found that risk for AF was increased by 25% with omega-3 treatment (1). Bae et al. reported similar associations (2), and Jai et al. found lower risk for AF among the five low dose omega-3 FA studies compared with risk in three higher risk studies (3).

Emerging evidence, however, has called these findings into question. For example, a 2023 biomarker-based meta-analysis of 17 prospective cohorts found that higher circulating levels of marine omega-3 FAs were inversely associated with incident AF suggesting a potential protective impact of low to intermediate doses known to produce such blood levels (4). Furthermore, methodological concerns have been raised regarding prior interventional analyses including inconsistent reporting of AF outcomes, potential informative censoring due to longer survival in omega-3 treated groups compared to placebo groups (5), and/or discrepancies in AF case ascertainment (6). These factors may have contributed to an overestimation of AF risk in prior analyses.

According to PubMed (8/16/2025), over the last 40 years there have been over 3900 published RCTs involving omega-3 fatty acids, thus there is significant opportunity to re-examine the omega-3/AF issue using a broader evidence base by including supplementation studies where AF was not a prespecified outcome but would have been recorded as an adverse event had it developed. The goal of this analysis was to conduct a more comprehensive review of the literature and to perform updated meta-analysis of RCTs to better understand the relationship between omega-3 supplementation and AF risk.

In contrast to earlier meta-analyses (1–3), we integrated both published and underutilized data sources, including trial registries and unpublished datasets, to reduce selective reporting bias. We further ensured that trials reporting no AF events contributed to the pooled evidence base. Finally, we applied a CVD risk/dose stratification framework to distinguish between doses largely achievable through diet or typical supplementation vs approved doses of pharmacological formulations.

## Methods

The protocol for this study is registered at the International Prospective Register of Systematic Reviews (**PROSPERO, CRD420251233813**). The conduct and reporting of this systematic review and meta-analysis adhered to the Preferred Reporting Items for Systematic Reviews and Meta-Analyses (PRISMA) 2020 guidelines (7). Ethical approval for this work was granted by the Human Ethics Board of the University of South Dakota (**IRB-21-136**). All individual RCTs included in this study received ethical approval from their respective institutional review boards or ethics committees, and all participants provided informed consent in the original studies.

### Eligibility criteria

To be eligible for inclusion in the meta-analysis, studies had to be RCTs investigating any omega-3 FA formulation containing eicosapentaenoic acid (EPA) and/or docosahexaenoic acid (DHA) administered in capsule form at a dosage that meets or exceeds the recommended daily intake of ≥500 mg/day endorsed by several international bodies (8–10). This followed the precedent of all prior meta-analyses which only included studies where EPA+DHA intakes exceeded this threshold. There was no upper limit on the dosage. Study designs were limited to parallel-group RCTs. Additional inclusion criteria included ≥12 months of treatment and the enrolment of participants ≥50 years of age. This criterion was based on the fact that risk for AF begins to accelerate after age 50 (11).

There were no restrictions on the relative proportions of EPA and DHA studied nor on the chemical form, i.e., ethyl esters, triglycerides (natural or re-esterified), monoacyl-glycerides, phospholipids, or carboxylic acids. RCTs administering EPA and/or DHA via liquid oil preparations (e.g., cod liver, seal, or tuna oil) were also eligible. Multi-component intervention studies (i.e., those combining omega-3 supplementation with pharmaceutical agents, antioxidant vitamins, or lifestyle interventions such as exercise regimens, low-fat diets and/or incorporated as part of a multi-nutrient supplement) were eligible if data from appropriate non-omega-3 control group were available.

We excluded trials that 1) administered omega-3 through dietary sources (e.g., oily fish meals), or through food/beverage-delivery based formats (e.g., fortified in margarine or a milkshake); 2) employed dietary regimen or lifestyle counselling as the sole intervention (e.g., Mediterranean diet) or advice to increase consumption of oily fish; 3) used omega-3 in the comparator arm (e.g., low vs high dose omega-3, or given dietary advice to increase oily fish intake, etc.); and 4) did not include a control/placebo or comparator group. Studies were also excluded if the intervention and control groups were not treated in parallel for at least 12 months, such as in delayed-start or open-label extension designs where the control group received the active intervention partway through the study.

We included studies regardless of participants’ health status. All patients with known AF (including paroxysmal, persistent, or permanent AF) at baseline were excluded from our analyses. For studies that enrolled participants both and without a history of AF, the trial was included, but data from participants with documented baseline AF were excluded from our dataset, where this was possible.

### Search strategy

A systematic search was conducted across nine databases: the Global Organization for EPA and DHA Omega-3s (GOED) Clinical Study Database, ResearchGate, PubMed, ClinicalTrials.gov, the International Standard Randomised Controlled Trial Number (ISRCTN) Registry, Australian New Zealand Clinical Trials Registry (ANZCTR), the National Institute for Health and Care Research (NIHR) Journal Library, the National Institutes of Health (NIH) Data Sharing and Archiving for Human Studies (DASH), medRxiv, Good Clinical Practice Network (GCP), and the Clinical Drug Experience Knowledgebase (CDEK). These databases were re-searched approximately monthly to detect newly published reports. The search was terminated on July 29, 2025. Search terms included different combinations of keywords related to the intervention and outcome of interest: ‘omega-3 fatty acid’, ‘fish oil’, ‘seal oil’, ‘cod-liver oil’, ‘tuna oil’, ‘algae oil’, ‘krill oil’, ‘calanus oil’, ‘icosapent ethyl (IPE)’, ‘DHA’, ‘EPA’, ‘lovaza’, ‘omega-3 acid ethyl esters (O3AEE)’, ‘vascepa’, ‘omacor’, ‘epanova’, ‘docosahexaenoic’, and/or ‘eicosapentaenoic’. Outcome terms included ‘incident atrial fibrillation,’ ‘new-onset atrial fibrillation’, and ‘atrial flutter.’ These terms were applied to titles and topic fields where database search functionality allowed. To ensure comprehensive coverage, reference lists of prior systematic reviews and key primary studies were also screened.

To minimize reporting bias, a targeted search strategy was developed to uncover RCTs that potentially collected relevant data but may not have been published in peer-reviewed journals. Using the same keywords listed above; pre-prints from medRxiv, and clinical trials registries (ANZCTR, ISRCTN, ClinicalTrials.gov, GCP); irrespective of trial status (i.e., on-going or completed) were all screened. Trials that were still recruiting, still in the intervention phase, terminated early, or had an unknown status were excluded. A targeted search of ResearchGate was conducted specifically to identify potentially relevant conference communications. Additionally, the NIH DASH repository was searched for publicly available datasets, and CDEK and Clinicaltrials.gov AEs records were queried specifically to review SAE data from RCTs investigating omega-3 products.

Titles and abstracts identified through the search were screened by one reviewer (NA), and all excluded records were independently checked by a second reviewer (WH). Full-text articles were assessed for eligibility by the two reviewers independently. Any disagreements at either stage were resolved through discussion and, when needed, consultation with a third reviewer (FQ). Screening was performed in an unblinded manner.

### Outcome

The primary outcome was the occurrence of new-onset AF as determined through central adjudication, ECG, electronic medical records (EMRs), insurance claims, and/or patient-reported adverse events.

### Data collection and extraction

One reviewer (NA) extracted all relevant data from included studies, and a second reviewer (WH) independently verified the accuracy and completeness of the extracted information. From each included study, the following information was abstracted: trial name, clinical trial registration number (if available), authors, year of publication, study design, comparator (placebo/control), omega-3 FA dosage and formulation, supplementation period, number of participants, patient characteristics, target population, and outcomes of interest. No attempt was made to standardize definitions of end points. Study quality was assessed using the modified Jadad scale (0-8 points). Scores of ≥5 indicated high-quality trials.

For this review, it was agreed that “published data” would be defined as reports of new-onset AF events extracted from original peer-reviewed publications or their supplementary materials. These events could be reported either as SAEs, pre-defined safety endpoints, or as pre-specified primary/secondary clinical outcomes of the study. Published data also included new-onset AF events reported in publicly accessible sources such as NIHR safety reports, ClinicalTrials.gov and CDEK SAE records, and systematic reviews or other secondary reports, even if these data were not in the original publications.

SAEs reported in published RCT articles, supplementary materials, ClinicalTrials.gov, and CDEK records were systematically reviewed to identify potential cases of new-onset AF. Events reported under broader or indirect headings such as ‘‘cardiovascular events,’’ ‘‘cardiac events,’’ ‘‘arrhythmic events,’’ ‘‘medication changes,’’ or ‘‘palpitations’’ were not considered equivalent to AF unless further clarification was obtained from investigators.

In trials where the protocol specified that adverse event monitoring followed the NIH Common Terminology Criteria for Adverse Events (12), which includes CV and rhythm-related events such as AF, the absence of reported AF was interpreted as absence of incident AF during follow-up. Likewise, when cardiac safety was a prespecified outcome and AE monitoring included systematic assessments (e.g., scheduled ECGs, or repeated cardiac evaluations), and investigators explicitly defined cardiac events but reported none as AF, these were accepted as valid findings. In such situations a value of zero AF events was entered for both trial arms.

For studies that did not prespecify AF as an outcome and did not explicitly report it under AEs or SAEs, we did not assume that there were zero events until confirmed by direct contact with principal investigators who were asked to review their AE datasets. If they confirmed that no AF events had been recorded, and if based on the study’s monitoring protocol incident AF would been reported if present, these studies were included and coded as zero events in both arms. Investigators were not asked to perform retrospective chart reviews or additional adjudication.

### Acquiring potential unpublished data

Many omega-3 studies outside of CVD have been conducted in individuals over the age of 50 who were given over 500 mg of EPA/DHA per day and followed for at least 12 months for study outcomes. We sought to obtain information about incident AF from these studies as well by proactively contacting PIs/corresponding authors directly and, where applicable, submitting formal data access request requests. If investigators responded with ‘no information available’, follow-up inquiries were sent to clarify whether this indicated that AF data were not collected, data had been collected but could not yet be shared, data status was unclear, AF events were not observed, or AF outcomes were not actively sought. In such cases, a lack of confirmation was not interpreted as zero AF events unless clearly stated and justified by the study’s monitoring procedures and confirmed by the PI. A maximum of three follow-up attempts were made to contact co-authors in cases of non-responding lead authors. All direct investigator outreach efforts were concluded in November 2025.

For trials without a published article, registry entries and/or study protocols were reviewed to determine whether new-onset AF was listed as a planned or monitored safety or clinical outcome (either explicitly or under broader categories such as arrhythmic or CV events).

Where such data were expected to have been collected but could not be located (e.g., because the trial remained unpublished, data under embargo, or in press), PIs were contacted to request access to the relevant data for this review (**Supplemental Table 2**).

### Data synthesis

Since CVD risk status and the EPA/DHA dose may each plausibly influence the development of AF, and this influence may be synergistic, we pre-specified analyses which simultaneously stratified by both variables to explore their potential modifying effects.

Stratification was applied at the trial level for CVD risk status and at the arm level for omega-3 dose. Accordingly, trials were classified into four strata: High Risk-High Dose (HR-HD), High Risk-Low Dose (HR-LD), Low Risk-High Dose (LR-HD), and Low Risk-Low Dose (LR-LD).

High risk for CV events was defined at the trial level as enrollment of participants with clinically diagnosed atherosclerotic cardiovascular disease (ASCVD) at baseline or having multiple (at least 2) CV risk factors (e.g., type-2 diabetes mellitus, hyperlipidemia, hypertension, recent myocardial infarction). Low risk for CV events was defined as participants without diagnosed ASCVD, including those enrolled for neurological, cognitive, autoimmune, or general wellness outcomes, or with one isolated CV risk factor (e.g., hypertension) but no confirmed CVD.

As noted, we stratified trials by omega-3 dose, distinguishing between levels achievable through fish consumption and/or use of dietary supplements vs those requiring pharmacological formulations, using a threshold of using a threshold of ≤ vs >1500 mg/day. This threshold was selected based on several factors: 1) it is below the lowest approved daily dose of pharmaceutical-grade omega-3 products (i.e., Epadel at 1800 mg/d) and 2) it has been shown to produce upper quintile levels of the omega-3 index [i.e., erythrocyte EPA+DHA, a validated biomarker of omega-3 intake (13, 14)] which have been associated with the lowest risk of AF (15), stroke (16), and all-cause mortality in large long-term epidemiological studies (17). Further details on the rationale used to distinguish EPA/DHA intakes from dietary supplements vs pharmacological agents can be found in

### Supplemental Appendix 1

For each study, AF event counts and total participant numbers were extracted separately for omega-3 intervention and control arms. Where trials had multiple omega-3 dose arms or multiple comparator arms, groups were pooled to avoid double-counting the comparator population. Pooling of omega-3 arms depended on whether all doses fell within the same predefined category. If all doses were in the same category (e.g., low dose), they were combined into a single treatment group, and the mean dose calculated. When a study investigated multiple omega-3 doses, each dose arm was stratified separately according to the high-dose vs low-dose criteria, and the non-omega-3 comparator group was necessarily included in both strata to allow for appropriate comparisons.

For all included trials, AF event counts and denominators were checked for completeness and internal consistency. When AF data were missing, incomplete, or unclear, study investigators were contacted to request clarification. If AF data remained unresolved after these inquiries, these RCTs were not included in the meta-analysis. However, studies with partially missing data were retained when analyzable arms were available.

### Data analysis

Absolute risk for AF was computed as the number of reported AF events divided by the number of subjects in the treatment group. The relative risk for AF was computed by dividing the absolute risk in the omega-3 group by that of the control group.

Logistic regression models, with Firth’s correction to adjust for small sample sizes, were fit on each cohort when predicting odds ratios (ORs). Cohort-specific ORs were then pooled by inverse-variance weighted, random effects meta-analysis using the metafor package in R. As a sensitivity analysis, we also conducted meta-analysis on the absolute risk difference using the metabin package in R. A second sensitivity analysis excluded studies which did not assess prevalent atrial fibrillation status at baseline. In all cases heterogeneity was assessed by the I^2^ statistic and further explored by meta-analyzing prespecified subgroups as created by classifying cohorts based on participants baseline CVD risk (high vs. low), and the omega-3 dose (> or < 1500 mg/day). A significance level of 0.05 was used for all analyses.

## Results

### Results of the search

**Figure 1** presents the study selection process and the acquisition of unpublished data, following the PRISMA 2020 guidelines (7). The electronic search yielded 3,688 records, of which 1,962 were duplicates and removed. Title and abstract screening of the remaining 1,726 records led to the exclusion of 1,647 studies. The primary reasons for exclusion were trial duration <12 months and/or mean participant age <50 years (n=1274).

**Figure 1.**
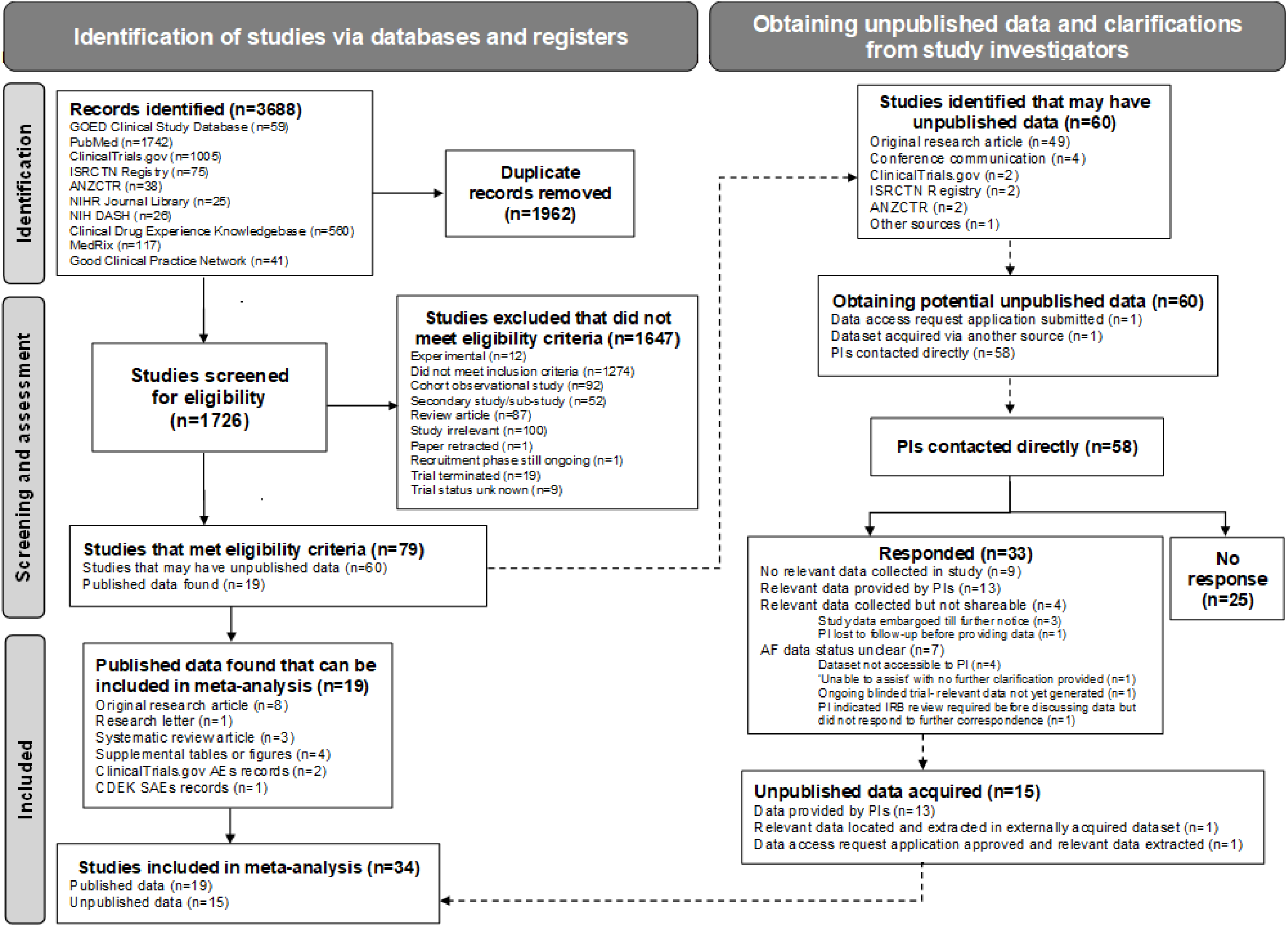
PRISMA 2020 flow diagram of study selection process and acquisition of unpublished data.

Additional reasons for exclusion are shown in **Figure 1**.

After full-text assessment of the remaining 79 RCTs that met the inclusion criteria; 19 had available published data on incident AF, while 60 were identified as studies potentially containing unpublished data. Formal data access requests were submitted where applicable (n=1; ADCS-DHA; NCT00440050), or PIs were contacted directly (n=58). One dataset (AREDS2; NCT00345176) was obtained via internal collaboration from a co-author (FQ) who had previously received the dataset.

Among the 58 PIs contacted, 33 responded. Of these, 13 provided usable AF data, 9 confirmed that AF data was not collected in their study, 4 confirmed relevant data collected but not shareable-either because it is currently embargoed (n=3; PreventE4; NCT03613844, PISCES; ISRCTN00691795, NUTRIMEMO; NCT02626247),or we were unable to obtain the data because no additional responses were received from PI after several follow-up attempts (n=1). 7 could not confirm status of AF data-either because raw datasets could not be accessed or reviewed and/or it remained unclear whether AF events were captured within broader AE categories (n=4); or because the trial was still ongoing and relevant safety data had not yet been generated (n=1; EMT2; NCT03428477); or because they responded that they were ‘unable to assist’ without confirming whether AF data existed (n=1); or indicated that institutional ethics approval would be required to clarify the availability of AF data but did not confirm whether such data had been collected and did not reply to further correspondence (n=1). Twenty-five PIs did not respond after three follow-up attempts (**Supplemental Table 2**).

In total, 34 RCTs were included in the meta-analysis. This included 19 RCTs with published AF data and 15 with unpublished AF data (13 obtained directly from PIs, one via a granted data request, and one through internal collaboration). Published data sources included original research articles (n=8), research letter (n=1), systematic review (18) (n=3), supplemental tables or figures (n=4), ClinicalTrials.gov (n=2) and CDEK (n=1) adverse event records.

### Characteristics of trials

Among the 34 RCTs there were 36 datasets comprising 114,326 participants (**Table 1**). In the pooled dataset, the mean baseline age was 66 years (range 53–78) with 62% men.

**Table 1.** Summary of 34 RCTs (36 datasets) including data from 114,374 subjects and the observed AF rates* segregated by high vs low dose EPA+DHA (< vs > 1500 mg/d) and by high vs low risk for CVD. Summary AF rates were based on weighted analysis by sample size.

| Trial/Authors (Year) | Omega-3 EPA/DHA |  |  | Comparator |  |  | Dosage | Duration |
| --- | --- | --- | --- | --- | --- | --- | --- | --- |
| *High dose >1500mg E+D/d* | AF Events | N | AF% | AF Events | N | AF% | EPA/DHA (mg/day) | Month |
| **High Risk High Dose (HR-HD)** |  |  |  |  |  |  |  |  |
| (54, 55) Yokote et al (2020)<sup>^</sup> | 0 | 306 | 0.0% | 0 | 77 | 0.0% | 2250 | 12 |
| (34, 35) REDUCE-IT (2023) | 103 | 3721 | 2.8% | 80 | 3707 | 2.2% | 4000 | 58.8 |
| (56) STRENGTH (2020) | 144 | 6539 | 2.2% | 86 | 6539 | 1.3% | 3000 | 42 |
| (57) FAAT (2005) | 3 | 164 | 1.8% | 3 | 164 | 1.8% | 2600 | 12 |
| (58) PEACH (2018)** | 0 | 73 | 0.0% | 0 | 144 | 0.0% | 1800 | 12 |
| (59) OMEMI (2023) | 28 | 387 | 7.2% | 15 | 372 | 4.0% | 1800 | 24 |
| (60) EVAPORATE (2020) | 0 | 31 | 0.0% | 0 | 37 | 0.0% | 4000 | 18 |
| (61) HEARTS (2025) | 8 | 111 | 7.2% | 5 | 102 | 4.9% | 3360 | 30 |
| (62) SAREPITASO (2018) | 0 | 38 | 0.0% | 0 | 38 | 0.0% | 1800 | 12 |
| (18, 63) Derosa et al (2016) | 1 | 128 | 0.8% | 3 | 130 | 2.3% | 2550 | 18 |
| (18, 64) OFAMI (2001) | 8 | 150 | 5.3% | 15 | 150 | 10.0% | 3360 | 18 |
| (37) RESPECT-EPA (2024) | 39 | 1244 | 3.1% | 20 | 1251 | 1.6% | 1800 | 60 |
| **Weighted** | **334** | **12892** | **2.6%** | **227** | **12711** | **1.8%** | **3109** | **46** |
| **High Risk Low Dose (HR-LD)** |  |  |  |  |  |  |  |  |
| (36) GISSI-HF (2013) | 278 | 2492 | 11.2% | 244 | 2499 | 9.8% | 870 | 46.8 |
| (65) Risk & Prevention (2013) | 113 | 6239 | 1.8% | 92 | 6266 | 1.5% | 925 | 60 |
| (38, 39) ASCEND (2018) | 596 | 7740 | 7.7% | 588 | 7740 | 7.6% | 840 | 88.8 |
| (18, 66) GISSI-Prevenzione (2001) | 40 | 2835 | 1.4% | 46 | 2828 | 1.6% | 860 | 42 |
| (67) SU.FOL.OM3 (2010) | 33 | 1253 | 2.6% | 32 | 1248 | 2.6% | 600 | 56.4 |
| (68) ORIGIN (2012) | 288 | 6281 | 4.6% | 259 | 6255 | 4.1% | 840 | 74.4 |
| **Weighted** | **1348** | **26840** | **5.0%** | **1261** | **26836** | **4.7%** | **854** | **68** |
| **Low Risk High Dose (LR-HD)** |  |  |  |  |  |  |  |  |
| (69) RCT-EPA (2023) | 0 | 61 | 0.0% | 0 | 60 | 0.0% | 3000 | 14 |
| (70) Mengelberg et al (2022) | 0 | 37 | 0.0% | 0 | 35 | 0.0% | 1842 | 12 |
| (71, 72) seAFOod Polyp Prevention (2018) | 3 | 347 | 0.9% | 0 | 350 | 0.0% | 2000 | 12 |
| (73, 74) EnRGISE (2019)<sup>#</sup> | 0 | 148 | 0.0% | 4 | 141 | 1.4% | 2100 | 12 |
| (75, 76) ADCS-DHA (2010) | 2 | 238 | 0.8% | 1 | 164 | 0.6% | 2040 | 18 |
| (77) Lin et al (2022)<sup>†</sup> | 0 | 40 | 0.0% | 0 | 40 | 0.0% | 1600 | 24 |
| (78) MARINA (2011)<sup>†</sup> | 0 | 92 | 0.0% | 0 | 88 | 0.0% | 1800 | 24 |
| (79) BRAVE-EPA (2025) | 0 | 63 | 0.0% | 2 | 68 | 2.9% | 4000 | 18 |
| (80) EPOCH (2018) | 2 | 195 | 1.0% | 1 | 196 | 0.5% | 2320 | 18 |
| (81) DREAM (2018) | 2 | 349 | 0.3% | 1 | 186 | 0.5% | 3000 | 12 |
| (82) CAPFISH-3 (2024) | 1 | 50 | 2.0% | 0 | 50 | 0.0% | 2200 | 12 |
| **Weighted** | **10** | **1620** | **0.6%** | **9** | **1378** | **0.7%** | **2355** | **15** |

| Low Risk<br>Low Dose<br>(LR-LD) |  |  |  |  |  |  |  |  |
| --- | --- | --- | --- | --- | --- | --- | --- | --- |
| (83, 84) Shinto et al (2014) | 1 | 34 | 2.9% | 0 | 33 | 0.0% | 1275 | 18 |
| (85) VITAL Rythm (2021)<sup>‡</sup> | 469 | 12542 | 3.7% | 431 | 12577 | 3.4% | 840 | 63.6 |
| (86) CANN (2023) | 0 | 125 | 0.0% | 0 | 121 | 0.0% | 1500 | 12 |
| (87, 88) AREDS2 (2013) | 242 | 2123 | 11.4% | 238 | 2056 | 11.7% | 1000 | 57.6 |
| (78) MARINA (2011)<sup>†</sup> | 0 | 187 | 0.0% | 0 | 88 | 0.0% | 675 | 24 |
| (77) Lin et al (2022)<sup>†</sup> | 0 | 83 | 0.0% | 0 | 40 | 0.0% | 925 | 24 |
| (89) DO-HEALTH (2025) | 34 | 1027 | 3.3% | 28 | 1037 | 2.7% | 990 | 36 |
| **Weighted** | **746** | **16121** | **4.6%** | **697** | **15928** | **4.4%** | **875** | **60** |

Abbreviations: AF: atrial fibrillation; CVD: cardiovascular disease; DHA: docosahexaenoic acid; EPA: eicosapentaenoic acid; N: number of participants randomized.
\*Details of AF ascertainment methods for each RCT are provided in **Supplemental Table 1**, and study characteristics are reported in **Supplemental Table 3**.
<sup>^</sup>Yokote et al. investigated two EPA doses (2g/day and 4g/day) in high-risk participants. Here we report the mean of the two doses.
<sup>\*\*</sup>PEACH investigated EPA 1.8 g/day administered with either 2 mg/day or 4 mg/day pitavastatin. As the EPA dose was identical across both statin strata, these two groups were combined in our analyses.
<sup>†</sup>MARINA and Lin et al., investigated multiple doses of omega-3 in low-risk participants. As a result, their low-dose arms were included in the LR-LD category, and their high-dose arms were included in the LR-HD category in the stratified analyses. We report the mean of the 2 low doses in MARINA (0.45g, 0.9g EPA+DHA) and Lin et al (0.7g DHA, 1150mg EPA+DHA).
<sup>‡</sup>VITAL-rhythm is an ancillary trial of the VITAL trial (NCT01169259)- our meta-analysis was performed using VITAL-rhythm data.
<sup>#</sup>EnRGISE participants received 1.4 g/day of fish oil for the first 6 months, after which the dose was escalated to 2.8 g/day for the subsequent 6 months. Here we report the average daily dose.

Daily omega-3 doses ranged from 600-4000 mg/day, and EPA/DHA was most commonly administered in either an ethyl ester (56%) or triglyceride-based formulation (32%). Most studies employed a double-blind, placebo-controlled design, with a smaller number conducted as open-label trials where usual care served as the control. A subset of trials adopted factorial or multi-factorial designs (44%), combining omega-3 supplementation with other active interventions. These included cardiometabolic drug therapies (e.g., statins, antiplatelet agents, antihypertensives, insulin), as well as vitamins (e.g., folate, vitamins B12, D or E) or structured lifestyle interventions such as physical activity or dietary counselling. Control arms included a wide variety of placebos ranging from oils (olive, corn, sunflower, soybean, mineral oil), with olive oil being the most employed placebo. The details of each individual study are presented in **Supplementary Table 3**.

### Rates of new-onset atrial fibrillation by dose/risk group

After stratifying the studies by dose and CVD risk status (**Table 1**), there were 12 studies in the HR-HD stratum including 25,603 patients given an average of 3109 mg EPA+DHA/day. The six HR-LD studies included 53,676 patients with an average dose of 854 mg/day. The LR-HD stratum included 11 studies with 2998 subjects given a mean dose of 2355 mg/d, and the LR-LD group included seven studies, 32,049 individuals with a mean dose of 875 mg/d.

### Meta-analysis

Looking at each of the four groups separately, only the HR-HD group (i.e., individuals at elevated CV risk receiving >1500 mg/day of EPA/DHA) showed a statistically significant increase in AF risk with a pooled odds ratio (OR) of 1.48; (**Figure 2**). The ORs for the other three categories were not statistically significant (1.07 for HR-LD; 0.95 for LR-HD and 1.06 for LR-LD). We also note that only two studies (both in the HR-HD group) found a statistically significant increase in risk of AF. In all four strata, residual heterogeneity estimates were less than 7% and not statistically significant (all four p>0.45). Across all studies, risk for AF was significantly increased by 14%.

**Figure 2.**
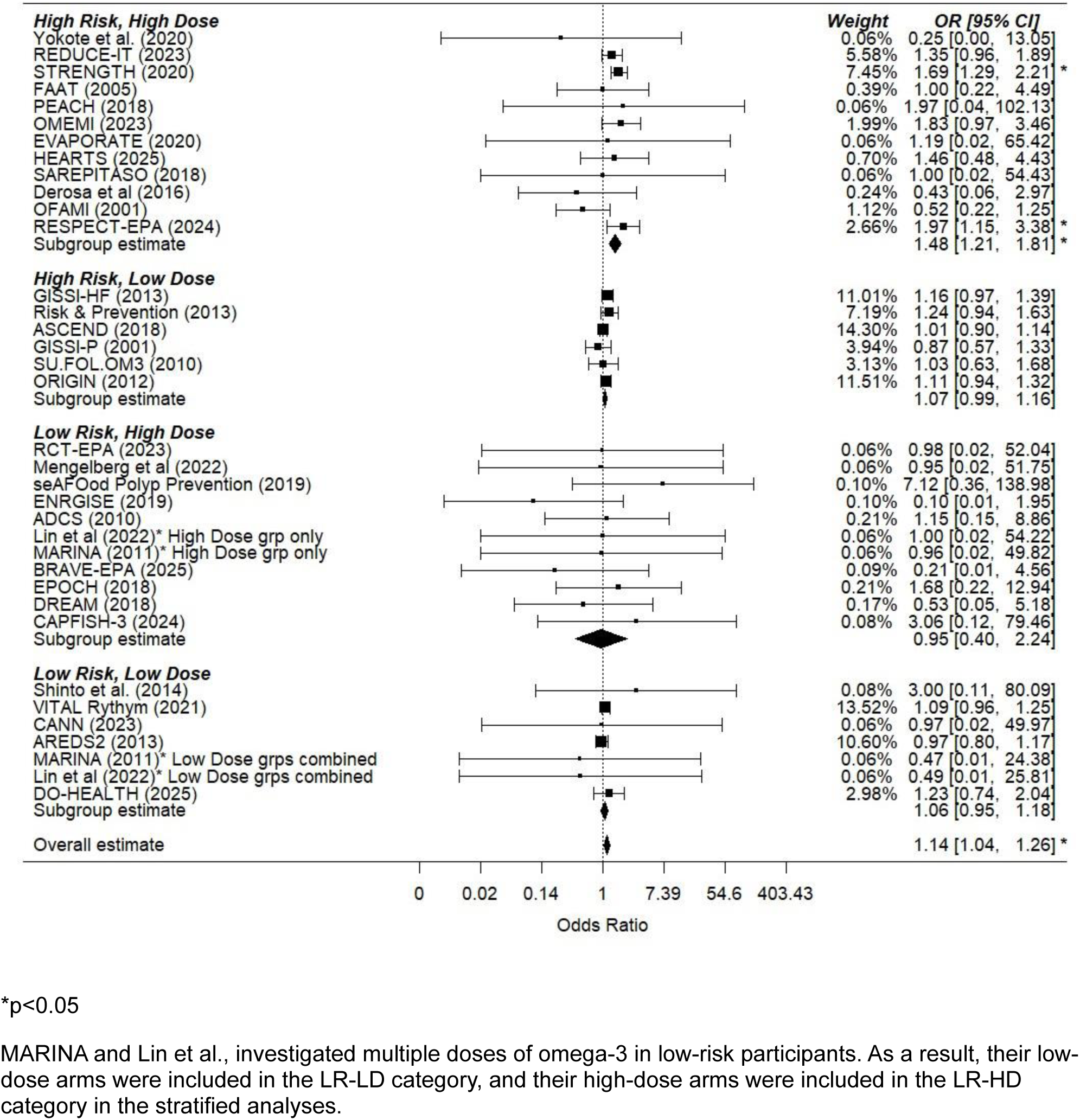
Meta-analysis of 34 RCTs including data from 114,374 subjects and the observed AF rates segregated by high vs low dose EPA+DHA (< vs > 1500 mg/d) and by high vs low risk for CVD.

Post hoc comparisons, using the HR-HD group as the reference, demonstrated the following (**Figure 2**). The HR-LD group exhibited a significantly lower AF risk (OR = 0.71, 95% CI: 0.59–0.87; p = 0.0007). Second, the LR-LD group had a significantly lower AF risk (OR = 0.71, 95% CI: 0.57–0.87; p = 0.0009). Finally, the LR-HD group showed the largest reduction in AF risk (OR = 0.63, 95% CI: 0.26–1.52) as compared to the HR-HD group, although this was not statistically significant (p = 0.31).

### Sensitivity analysis: Risk difference

**Supplemental Figure 1** illustrates a similar pattern of results when considering the risk difference (RDs) instead of ORs. Namely, only the HR-HD group showed statistically significant increased risk (RD = 0.8%, 95% CI: 0.40-1.1, p<0.0001).

**Supplemental Figure 2** illustrates a similar pattern of results when excluding studies which did not assess AF status at baseline (no prevalent AF). In three of four cases, the ORs were very similar to that obtained when considering all studies (HR-HD: OR 1.52 [no prevalent AF] vs. 1.48 [all studies]; HR-LD: OR 1.07 [no prevalent AF] vs. 1.07 [all studies]; LR-LD: OR 1.06 [no prevalent AF] vs. 1.06 [all studies]). Also similarly, the HR-HD group was the only one which was statistically significant. The LR-HD group OR was 0.54 in the no prevalent AF studies, and 0.95 for all studies-however, the confidence intervals for these estimates were very wide, and neither was statistically significant.

## Discussion

Recent meta-analyses reporting that DHA and EPA increase risk of AF (1–3) have raised doubts among physicians and the public about the safety of omega-3 products, whether dietary supplements or pharmaceutical preparations. As noted earlier, these previous meta-analyses included no more than eight RCTs whereas the present analysis now includes 34 RCTs. By examining how both background CV risk and omega-3 dose influence AF outcomes, our findings provide a more nuanced perspective on the relationship between omega-3 FAs and risk for AF.

We found a statistically significant, 48% increase in the relative risk of AF and an absolute risk increase of <1% in the HR-HD group. In contrast, the HR-LD, LR-HD, and LR-LD strata (**Figure 2**) showed pooled risk estimates that were substantially lower than that of the HR-HD group (ORs ∼1.0) and none were statistically significant. Importantly, post-hoc comparisons using HR-HD as the reference, HR-LD and LR-LD groups each showed approximately null and significantly lower AF risk (OR ≈ 0.70).

Among LR individuals, the mean intake in the HD group was approximately 2300 mg/day (**Table 1**), and this intake was associated with a reduced AF risk (OR = 0.67) as compared to the HR-HD group, and an OR<1 (OR=0.95) overall. This suggests that in the general population, EPA/DHA intakes at this level are safe to consume (vis-à-vis risk for AF). This finding, however, was based on relatively few events, therefore should be considered tentative. Nevertheless, this is consistent with the Food and Drug Administration’s designation of EPA/DHA intakes of up to 3000 mg/d are ‘generally recognized as safe’ (19).

There are a variety of other considerations that would support the safety vis-à-vis AF in the general population. For example, on average, North American adults eat <1 serving per week of fish/seafood; thus, the mean intake of DHA+EPA in the United States (US) is only ∼100 mg/d (20). This is associated with a mean DHA+EPA level in red blood cells [i.e., the omega-3 index (13)] of around 5.5 % in the US (21). Observational studies consistently show that an omega-3 index of 8% or higher is ideal for reducing risk of MACE (22), stroke (16) and all-cause mortality (23). For typical Americans, achieving this would require an increased consumption of about 1000 mg/d of DHA+EPA (14). The Japanese consume about this amount (24) and notably have an age-adjusted incidence of AF that is about one-third that of the US population (25, 26). These ecological considerations are at least consistent with the view that in the general population, even relatively high doses (1-1.5 g/d) do not increase risk, and indeed may decrease risk, for AF.

There are several potential mechanisms that might explain the increased risk for AF in HR patients taking high-dose omega-3. Omega-3 treatment has been shown to reduce resting heart rate (27) through increasing vagal tone even at relatively low doses of about 500 mg/d (27, 28). These dose-dependent vagotonic actions of omega-3 suggest a mechanism that may in part explain its divergent effects on risk of developing AF, whereby low dose omega-3 does not significantly increase AF risk and may actually reduce risk, but high-dose omega-3 can increase AF, especially in high-risk individuals (29). These patients also often have structural heart disease and many of them are on other medications such as beta blockers, which can also block sympathetic activation and therein increase vagal tone. The dose-dependent vagal-stimulating effects are likely to be especially problematic for individuals at high risk for bradycardia-dependent AF (30). This high-risk for AF includes individuals who perform large quantities of strenuous endurance exercise, have untreated obstructive sleep apnea, or sick sinus syndrome and chronically slow heart rates (30). A re-analysis of data from high-dose omega-3 RCTs that explored interactions with baseline heart rate, omega-3 intake and AF incidence would help to clarify this potential relationship.

Considering that stroke is the major complication of AF, it is paradoxical that higher omega-3 intake and blood levels have been repeatedly associated with a lower risk of stroke. Specifically, a comprehensive meta-analysis (n > 180,000 subjects) found patients in the top quintile of marine omega-3 blood levels had an 18% lower risk of ischemic stroke, and no effect on hemorrhagic stroke compared to patients in the lowest quintile of omega-3 (16). Similarly, in the REDUCE-IT trial, which was comprised of high-CVD risk patients, high-dose EPA (4,000 mg/d) reduced relative risk of stroke by 28% despite increasing relative risk of AF by 35% (31). Even those in the EPA arm of the REDUCE-IT study who developed AF experienced lower risks of stroke and MACE compared to those in the control group who developed AF (31). This presumed protective effect against stroke may relate to the mild anti-thrombotic effects of EPA and DHA (32, 33). Still, for high-risk people with a history of AF, Olshansky et al. reported a non-significantly higher risk for developing AF on EPA than similar patients in REDUCE-IT without a history of AF (34). Accordingly, a lower omega-3 target dose of <1500 mg/d of DHA/EPA may be a safer level of intake for CVD patients with a history of AF.

The potential risk of AF must be considered within the broader risk–benefit profile of omega-3 therapy, particularly in HR populations where higher doses are typically used. Across several major RCTs-such as REDUCE-IT (34, 35), GISSI-HF (36), RESPECT-EPA (37), and ASCEND (38, 39); the magnitude of cardiovascular benefit equaled or exceeded the observed increase in AF risk. For example, in REDUCE-IT, the 1.0% absolute increase in AF hospitalization with 4 g/day EPA was offset by a 4.8% absolute reduction in major adverse cardiovascular events (MACE), a trade-off that strongly favors treatment given that events such as myocardial infarction, ischemic stroke, or cardiovascular death carry far greater clinical consequences than increased risk for new-onset AF. This interpretation aligns with recent cost-effectiveness analyses demonstrating that the reduction in MACE more than compensates for increased AF risk, supporting an overall favorable benefit–risk balance for high-dose omega-3 therapy (40).

Meta-analyses of prospective cohort studies focusing on omega-3 blood levels as the exposure have reported lower risk for mortality from all causes and from CVD, cancer and remaining other causes (17, 22). Other biomarker-based studies have found higher levels of omega-3s to be associated with lower risk for ASCVD (23), Alzheimer’s disease (41) and total dementia (41), heart failure (42), diabetes (43), suicidal ideation/self-harm (44), poor lung function (45), colorectal cancer and several other malignancies (46, 47), atopic dermatitis (48), hospitalization for sepsis (49), liver cirrhosis (50), chronic kidney disease (51), frailty (52) and hip fractures (53). These associations, in addition to a lower risk for AF (4), support a generalized overall benefit of omega-3 in human health.

### Strengths and Limitations

This meta-analysis represents a substantial advance in our understanding of the relationship between omega-3 treatment and risk for AF. Unlike earlier work that relied almost exclusively on published reports, we systematically integrated underutilized data sources, including trial registry adverse event records and unpublished datasets; thereby reducing selective reporting bias. Importantly, we also incorporated trials in which no AF events were observed (where, had they occurred, they would have been noted), ensuring that such data contributed to the overall evidence base rather than being excluded. A further novel contribution is our risk-dose stratification framework which moves one step beyond the ‘high dose vs low dose’ paradigm used in past studies (1). This approach provides a clearer understanding of how simple dietary supplementation might affect risk for AF as opposed to the use of high-dose pharmaceutical products. It also illuminates the importance of patient substrate, as we found limited evidence of an increase in AF risk even in high-risk CVD patients who are taking relatively low omega-3 doses (<1500 mg/d). Finally, by including populations outside of high-risk CV cohorts, our analysis broadens generalizability and offers more nuanced insight into how AF risk may differ across clinical contexts. There are also limitations to this work, chief among them is the admitted inability to ensure that all relevant studies - published or not - were included in the analysis. Having made our best effort to identify relevant data sets, there were several studies published decades ago that could not be included as contact with the original authors proved impossible.

## Conclusions

This meta-analysis revealed that high doses of omega-3 fatty acids (typically from pharmacological products) given to patients at high risk for ASCVD increased the relative risk for AF by approximately 50%, though the absolute risk increase is modest (0.8%). In all other settings (HR-LD, LR-HD, and LR-LD) there was no statistically significant increase in risk for AF with omega-3 supplementation. Thus, fish oil supplementation providing less than 1500 mg of EPA+DHA per day (and possibly up to 2400 mg) does not appear to increase risk for AF. On the other hand, doses of >3000 mg/d in high-risk CVD patients may warrant caution. Given that elevated blood omega-3 levels (driven by both dietary choices and supplementation) are associated with reduced risk for ASCVD (and other health outcomes), the small AF signal in HR-HD populations should be weighed against substantial CVD benefit when making treatment decisions. New RCTs specifically designed to evaluate the risk:benefit ratio of HD omega-3 supplementation in HR populations should be undertaken.

## Data Availability

All data produced in the present work are contained in the manuscript.

https://www.adcs.org/

## Disclosures

JHO is the Chief Medical Officer of CardioTabs, a company that sells Omega-3 products. WSH holds stock in OmegaQuant Analytics, LLC, a laboratory which offers blood fatty acid testing (including the Omega-3 Index) for researchers, clinicians, and consumers. None of the other authors have any conflicts to disclose.

## Abbreviations

AF: Atrial Fibrillation
CV: Cardiovascular
CVD: Cardiovascular Disease
DHA: Docosahexaenoic Acid
EPA: Eicosapentaenoic Acid
ISSFAL: International Society for the Study of Fatty Acids and Lipids
PIs: Principal Investigators
RCTs: Randomized Controlled Trials
IPE: Icosapent Ethyl
O3AEE: Omega-3 Acid Ethyl Esters
OCA: Omega-3 Carboxylic Acids
HR-HD: High Risk-High Dose
HR-LD: High Risk–Low Dose
LR-HD: Low Risk–High Dose
LR-LD: Low Risk–Low Dose
MA: Meta-analysis

## Supplementary materials

**Supplementary Appendix 1-** Rationale used to distinguish EPA/DHA intakes from dietary supplements vs pharmacological agents.

**Supplementary Appendix 2-** Funding statements for RCTs included in MA.

**Supplemental Table 1**- RCTs included in MA with AF ascertainment method and data publication status.

**Supplemental Table 2**- RCTs meeting eligibility criteria but excluded from MA.

**Supplemental Table 3**- Study characteristics of RCTs included in MA.

**Supplemental Figure 1**- Risk differences sensitivity analysis of all RCTs included in MA.

**Supplemental Figure 2**- Odds ratio sensitivity analysis including only RCTs that explicitly excluded individuals with a history of AF at enrollment or prevalent AF at baseline.

## Author contributions

NA: developed search strategy, led literature searches and study selection, coordinated investigator outreach and data acquisition, performed data extraction and curation, drafted and revised the manuscript; NLT: statistical analysis and critical review; WSH: project oversight and critical review; FQ, YL, YS, HQ: contributed to data collection and critical review; JHO: critical review; all other authors provided study specific data and critically reviewed the manuscript.

## Funding

The author(s) declare that financial support was received for the research and publication of this article. The project was supported by The Global Organization of Omega-3 EPA and DHA (GOED) which had no role in the research design or conduct; data analysis or interpretation; preparation, review, or approval of the manuscript; or the decision to submit the manuscript for publication. FQ is supported by the T32HL125232 Multidisciplinary Training Program in Cardiovascular Epidemiology.

## Acknowledgments

We thank Hertzel C. Gerstein MD MSc FRCPC, for providing access to data from the ORIGIN trial (NCT00069784). Data from the ADCS-DHA trial (NCT00440050) used in the preparation of this manuscript were obtained from the University of California, San Diego Alzheimer’s Disease Cooperative Study (ADCS) legacy database (https://www.adcs.org/). Data collection and sharing for this project were funded by the ADCS (National Institutes of Health Grant U19 AG010483). Data from the AREDS2 trial (NCT00345176) were obtained from the National Eye Institute, National Institutes of Health and is available through dbGaP (accession phs002015.v2. p1). Funding sources for all other included RCTs are provided in **Supplemental Appendix 2**.

## Notes

### Clinical Protocols

https://www.crd.york.ac.uk/PROSPERO/view/CRD420251233813

### Author Declarations

Ethical approval for this work was granted by the Human Ethics Board of the University of South Dakota (IRB-21-136). All individual randomized-controlled trials included in this meta-analysis received ethical approval from their respective institutional review boards or ethics committees, and all participants provided informed consent in the original studies.

