## Supplemental Figures 1-2 for "Effects of Omega-3 Fatty Acid Treatment on Risk for Atrial Fibrillation: An Updated Meta-Analysis of 34 Trials including 114,326 Individuals"

**Supplemental Figure 1.** Risk differences (RD) sensitivity analysis of 34 RCTs including data from 114,374 participants, and the observed AF rates segregated by high vs low dose EPA+DHA (< vs > 1500 mg/d) and by high vs low risk for CVD.

| Study | Weight | RD | 95%-CI |
|---|---|---|---|
| **High Risk, High Dose** | | | |
| Yokote et al. (2020) | 1.0% | 0.000 | [-0.018; 0.018] |
| REDUCE-IT (2023) | 8.6% | 0.006 | [-0.001; 0.012] |
| STRENGTH (2020) | 16.4% | 0.009 | [ 0.004; 0.013] |
| FAAT (2005) | 0.4% | 0.000 | [-0.029; 0.029] |
| PEACH (2018) | 0.8% | 0.000 | [-0.021; 0.021] |
| OMEMI (2023) | 0.3% | 0.032 | [-0.001; 0.065] |
| EVAPORATE (2020) | 0.1% | 0.000 | [-0.056; 0.056] |
| HEARTS (2025) | 0.1% | 0.023 | [-0.041; 0.087] |
| SAREPITASO (2018) | 0.1% | 0.000 | [-0.050; 0.050] |
| Derosa et al (2016) | 0.4% | -0.015 | [-0.045; 0.015] |
| OFAMI (2001) | 0.1% | -0.047 | [-0.107; 0.013] |
| RESPECT-EPA (2024) | 2.3% | 0.015 | [ 0.003; 0.027] |
| **Subgroup effect** | **30.5%** | **0.008** | **[ 0.004; 0.011]** |
| **High Risk, Low Dose** | | | |
| GISSI-HF (2013) | 1.2% | 0.014 | [-0.003; 0.031] |
| Risk & Prevention (2013) | 16.7% | 0.003 | [-0.001; 0.008] |
| ASCEND (2018) | 4.7% | 0.001 | [-0.007; 0.009] |
| GISSI-P (2001) | 8.2% | -0.002 | [-0.009; 0.004] |
| SU.FOL.OM3 (2010) | 2.1% | 0.001 | [-0.012; 0.013] |
| ORIGIN (2012) | 6.5% | 0.004 | [-0.003; 0.012] |
| **Subgroup effect** | **39.4%** | **0.002** | **[-0.001; 0.005]** |
| **Low Risk, High Dose** | | | |
| RCT-EPA (2023) | 0.3% | 0.000 | [-0.032; 0.032] |
| Mengelberg et al (2022) | 0.1% | 0.000 | [-0.053; 0.053] |
| seAFOod Polyp Prevention (2019) | 2.6% | 0.009 | [-0.003; 0.020] |
| ENRGISE (2019) | 0.4% | -0.028 | [-0.059; 0.002] |
| ADCS (2010) | 1.2% | 0.002 | [-0.014; 0.019] |
| Lin et al (2022)* High Dose grp only | 0.1% | 0.000 | [-0.048; 0.048] |
| MARINA (2011)* High Dose grp only | 0.7% | 0.000 | [-0.021; 0.021] |
| BRAVE-EPA (2025) | 0.1% | -0.029 | [-0.078; 0.020] |
| EPOCH (2018) | 1.1% | 0.005 | [-0.012; 0.022] |
| DREAM (2018) | 2.3% | -0.003 | [-0.014; 0.009] |
| CAPFISH-3 (2024) | 0.1% | 0.020 | [-0.034; 0.074] |
| **Subgroup effect** | **9.2%** | **0.001** | **[-0.005; 0.007]** |
| **Low Risk, Low Dose** | | | |
| Shinto et al. (2014) | 0.1% | 0.029 | [-0.049; 0.108] |
| VITAL Rythym (2021) | 15.7% | 0.003 | [-0.001; 0.008] |
| CANN (2023) | 1.3% | 0.000 | [-0.016; 0.016] |
| AREDS2 (2013) | 0.9% | -0.003 | [-0.023; 0.016] |
| MARINA (2011)* Low Dose grps combined | 1.1% | 0.000 | [-0.017; 0.017] |
| Lin et al (2022)* Low Dose grps combined | 0.2% | 0.000 | [-0.037; 0.037] |
| DO-HEALTH (2025) | 1.5% | 0.006 | [-0.009; 0.021] |
| **Subgroup effect** | **20.8%** | **0.003** | **[-0.001; 0.007]** |
| **Overall effect** | **100.0%** | **0.004** | **[ 0.002; 0.006]** |

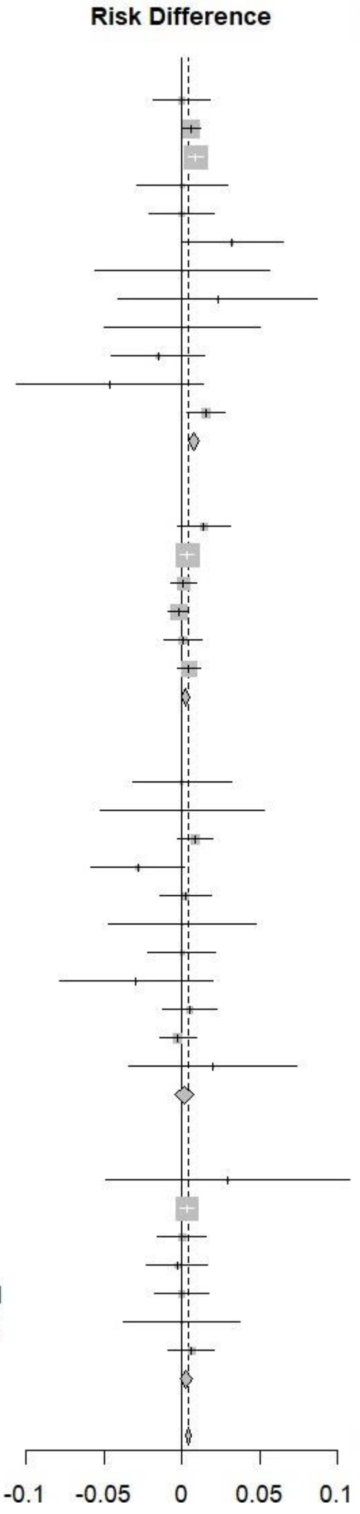

**Supplemental Figure 2.** Sensitivity analysis of the odds ratio (OR) of 15 RCTs (65,067 participants) that explicitly excluded individuals with a history of/prevalent AF at baseline. Observed AF rates segregated by high vs low dose EPA+DHA (< vs > 1500 mg/d) and by high vs low risk for CVD.

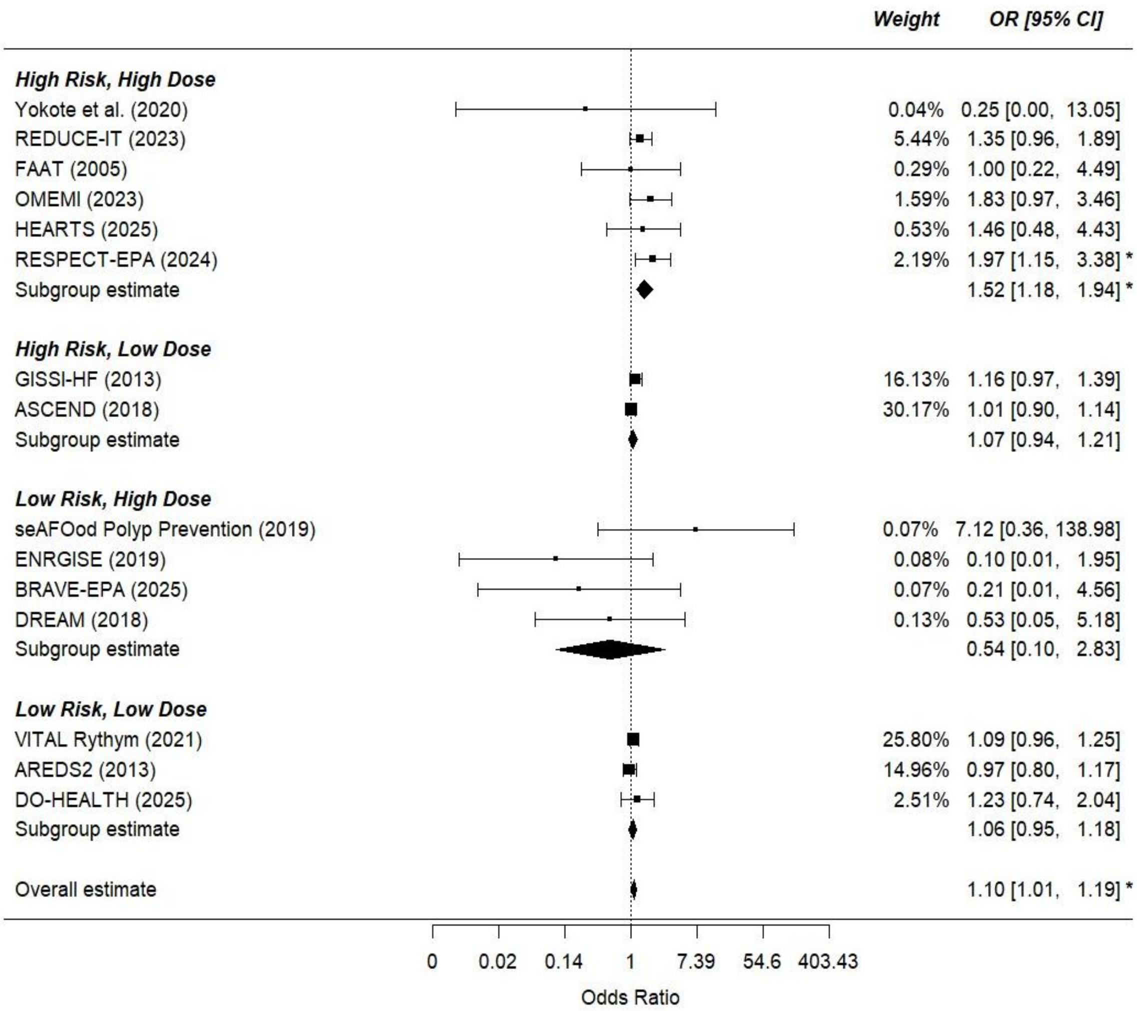


HEARTS, OFAMI, OMEMI, FAAT, RESPECT-EPA, DO-HEALTH, VITAL, REDUCE-IT, and AREDS2 enrolled mixed populations (including participants with AF at baseline) but reported baseline AF status, allowing exclusion of those with prior AF from all analyses.

GISSI-HF conducted baseline ECGs to ascertain AF and assessed history of paroxysmal AF.

ASCEND indirectly excluded known AF through anticoagulant exclusion criteria, later validated by electronic health record linkage.

MARINA confirmed the absence of AF by 24hr heart rate monitoring.

DREAM, ENrGISE and EVAPORATE excluded participants with prior or persistent/paroxysmal AF.

Yokote et al., BRAVE-EPA and SeaFOod Polyp Prevention excluded participants receiving anticoagulant therapy, effectively excluding those with prior AF.
