## Supplemental Table 1 for "Effects of Omega-3 Fatty Acid Treatment on Risk for Atrial Fibrillation: An Updated Meta-Analysis of 34 Trials including 114,326 Individuals"

**Supplemental Table 1.** 34 RCTs meeting eligibility and included in meta-analysis stratified by cardiovascular risk and omega-3 dosage (high dose >1500 mg E+D/d), with publication status, AF data source, and method of AF ascertainment**.**

| Trial/ Authors (Year) | Trial Full-name<br><br>Trial Registration No. (if applicable) | Publication Status<br><br>Data Source | Method of AF Ascertainment |
|---|---|---|---|
| **High Risk High Dose (HR-HD)** | | | |
| **Yokote et al (2020)**[1, 2] | AZD0585 Phase III Long-term Study in Japan<br><br>NCT02463071 | Published data<br><br>Data extracted from Clinical Drug Experience Knowledgebase AEs Records[2]. | AF not a prespecified outcome of Yokote et al, however, safety was a primary outcome of the study. Safety monitoring included AEs, vitals, labs, physical exams, and ECGs at 4-5 visits over 12 months. Serious cardiac events (e.g., MI, AV block) reported[2], but no AF events. Given the thorough cardiac monitoring design, AF considered absent by us (0 events). |
| **REDUCE-IT (2023)**[3,4, 5] | Reduction of Cardiovascular Events with Icosapent Ethyl-Intervention Trial<br><br>A Study of AMR101 to Evaluate Its Ability to Reduce Cardiovascular Events in High-Risk Patients With Hypertriglyceridemia and on Statin (REDUCE-IT)<br><br>NCT014923615 | Published data.<br><br>Data reported in research article[3] and clinicaltrials.gov AEs records[5] | AF and atrial flutter reported as a prespecified exploratory tertiary endpoints in REDUCE-IT, adjudicated by the Clinical Endpoint Committee. AF/atrial flutter hospitalizations (≥24 hours) were captured under 'arrhythmia requiring hospitalization.' Additional AF events not meeting adjudication criteria were captured through standard AE reporting. Annual ECGs were performed. AEs were coded using MedDRA v20.1[6] and included all treatment-emergent events from first dose until 30 days post-treatment.<br><br>Although post-hoc subgroup analyses by prior AF status have been published[3,] AE-level AF data restricted to participants without baseline AF were not available to our knowledge. Contact with PI was attempted to clarify this but received no response. |

| | | | |
|---|---|---|---|
| **STRENGTH (2020)[7]** | Outcomes Study to Assess STatin Residual Risk Reduction With EpaNova in HiGh CV Risk PatienTs With Hypertriglyceridemia (STRENGTH)<br><br>NCT02104817 | Published data.<br><br>Data reported in supplemental figure[7] | AF was a prespecified tertiary efficacy endpoint STRENGTH, reported as Kaplan–Meier curves for investigator-reported AF events. All events were adjudicated centrally by a core laboratory (C5Research). |
| **FAAT (2005)[8]** | Fatty Acid Antiarrhythmia Trial<br><br>NCT00004559 | Unpublished data.<br><br>Data acquired directly from PIs. | AF not a prespecified outcome of FAAT. PIs confirmed that patients were monitored for development of other arrhythmias (e.g. AF), requiring therapy captured via CRF medication-change field throughout the study period. PIs confirmed by reviewing their AEs records that these were due to new-onset AF development. However, events are not separately reported; thus, AF incidence is inferred rather than directly adjudicated. |
| **PEACH (2018)[9]** | The PEACH (Pitavastatin And Combined Omega-3 Health) Study<br><br>UMIN000003171. | Published data.<br><br>Data reported in research article[9]. | AF not a prespecified outcome of PEACH. Safety was assessed by recording all AEs and CV events of special interest (including arrhythmias). These were reported by site investigators to the PI, and then to the DSMB. No CV events were observed or reported during the study period; AF therefore, considered absent by us (0 events). |
| **OMEMI (2023)[10]** | OMega-3 fatty acids in Elderly patients with Myocardial Infarction study<br><br>NCT01841944 | Published data.<br><br>Data reported in research article[10]. | AF reported as a predefined secondary safety outcome in OMEMI. AF and micro-AF was detected by clinical assessment and by thumb-ECG screening, adjudicated by blinded investigators. Safety/event ascertainment was via both study visits and ECG monitoring. |
| **EVAPORATE (2020)[11]** | Effect of Vascepa (Icosapent Ethyl) on Improving Coronary Atherosclerosis in People with High Triglycerides Taking Statin Therapy (EVAPORATE)<br><br>NCT02926027 | Unpublished data.<br><br>Data acquired directly from PI. | AF reported as a prespecified safety outcome of EVAPORATE. PI confirmed that systematic ascertainment of AF conducted via ECGs, patient reports, physical exams at 3, 9, 12, 18 mos and hospital/clinic records. PI confirmed no AF events occurred in either group. |

| | | | |
|---|---|---|---|
| **HEARTS (2025)**[12] | Slowing HEART diSease With Lifestyle and Omega-3 Fatty Acids<br><br>NCT01624727 | Unpublished data.<br><br>Data acquired directly from PI. | AF was a prespecified safety endpoint. PI confirmed that cardiac events, including first occurrence of AF or atrial flutter, were monitored every 3 mos during the 30-month intervention. Medical records were obtained to confirm all events. |
| **SAREPITASO (2018)**[13] | SARpogrelate-EPA-PITavastatin-ASO Study | Published data.<br><br>Data reported in research article[13]. | AF not a prespecified outcome of SAREPITASO. CV events were monitored throughout the study and PIs explicitly report none were observed. AF therefore, considered absent by us (0 events). |
| **Derosa et al (2016)**[14] | Effects of n-3 pufas on fasting plasma glucose and insulin resistance in patients with impaired fasting glucose or impaired glucose tolerance | Published data.<br><br>Data extracted from systematic review[15] | AF not a prespecified outcome of Derosa et al (2016). Specific AF ascertainment details were not directly reported. Contact with PI was attempted to clarify this but received no response. |
| **OFAMI (2001)**[16] | Effects of High-dose n-3 Fatty Acids on Clinical Outcome and Serum Lipids – Omacor Following Acute Myocardial Infarction (OFAMI)<br><br>NCT01422317 | Published data.<br><br>Data extracted from systematic review[15] | AF not a prespecified outcome of OFAMI. The method of AF ascertainment is unclear; however, the primary publication[16] reported that repeated ECG recordings at 6 wks, 6-, 12- and 18 mos, and for some patients at 24 mos post-randomization were conducted. AF detection may therefore, have been captured as part of safety monitoring, but this was not explicitly reported. Contact with PI was attempted to clarify this but received no response. |
| **RESPECT-EPA (2024)**[17] | Randomized Trial for Evaluation in Secondary Prevention Efficacy of Combination Therapy-Statin and Eicosapentaenoic Acid<br><br>UMIN000012069 | Published data.<br><br>Data extracted from supplemental table[17] | AF was a prespecified secondary endpoint, captured under 'cardiac disease-related events.' AF events were systematically monitored at yearly visits for 3 yrs by ECGs during the study period, and recorded under both clinical outcomes and AEs reporting. All AF events were adjudicated by an independent endpoint committee blinded to treatment assignment. |

| | | | |
|---|---|---|---|
| **High Risk Low Dose (HR-LD)** | | | |
| **GISSI-HF (2013)[18]** | Gruppo Italiano per lo Studio della Sopravvivenza nell'Insufficienza Cardiaca (Italian Group for the Study of the Survival in Heart Failure)<br><br>GISSI-HF- Effects of n-3 PUFA and Rosuvastatin on Mortality-Morbidity of Patients With Symptomatic CHF<br><br>NCT00336336 | Published data.<br><br>Data reported in research article[18]. | AF reported as a predefined secondary outcome in GISSI-HF. Outcomes were collected from ECGs performed at each visit, and from events between visits (e.g., AF causing/worsening HF, AF causing hospitalization or as an event occurring during hospitalization), hospitalization records, or clinical documentation. |
| **Risk & Prevention (2013)[19]** | Risk and Prevention Study: Evaluation of the Efficacy of n-3 PUFA in Subjects at High Cardiovascular Risk<br><br>NCT00317707 | Published data.<br><br>Data extracted from supplemental table[19]. | AF reported as a primary efficacy endpoint in Risk & Prevention. This was captured under 'hospital admission for cardiovascular causes' (ICD-9 codes 390–459 or procedures 35–39). Events were documented with narrative summaries, supporting documentation, and adjudicated by a blinded committee of a cardiologist, internist, and neurologist. Only AF hospitalizations are reported; not clear whether AE-level AF data was collected. Contact with PI was attempted to clarify this, but no response was received. |
| **ASCEND (2018)[20, 21]** | ASCEND: A Study of Cardiovascular Events iN Diabetes<br><br>NCT00135226 | Published data.<br><br>Data reported in research letter[21]. | AF reported as an exploratory vascular outcome in ASCEND, but was not adjudicated clinically. Outcomes ascertained via hospitalizations, SAEs reports, ICD-10/OPCS codes, electronic record linkage to hospital episodes (covering 97% of participants with 14-yr look-back), and patient self-reports. |

| | | | |
|---|---|---|---|
| **GISSI-Prevenzione (2001)[22]** | Gruppo Italiano per lo Studio della Sopravvivenza nell'Infarto Miocardico –<br><br>Prevenzione (Italian Group for the Study of the Survival of Myocardial Infarction – Prevention) | Published data.<br><br>Data extracted from systematic review[15] | AF not a prespecified outcome of GISSI-Prevenzione. Specific AF ascertainment details were not directly reported in original research article[22]. Contact with PI was attempted to clarify this but received no response.<br><br>In GISSI-Prevenzione, only AF data from the n-3 alone and control groups are included in our dataset; AF data from other multi-factorial groups (vitamin E alone, n=2,830; n-3 + vitamin E, n=5,660) were not included in the systematic review[15]. Contact with PI was attempted to request access to this dataset but received no response. |
| **SU.FOL.OM3 (2010)[23]** | SUpplementation with FOlate, vitamin B6 and B12 and/or OMega-3 fatty acids<br><br>ISRCTN41926726 | Unpublished data.<br><br>Data acquired directly from PI. | AF was a prespecified secondary endpoint of SU.FOL.OM3 the trial. According to study protocol[24] participants attended annual in-person follow-up visits (T1, T2, T3, T5) during which physical examinations and biological assessments were performed across 257 study centers. PI confirmed AF was ascertained and recorded by ECG. Between annual visits, participants received semi-annual mailed questionnaires inquiring about study outcomes (including new diagnoses and hospitalizations). Non-responders were contacted and interviewed by study physicians via telephone.<br><br>Incident CV events (including AF) were further evaluated by a centralized adjudication process. Research assistants verified reported events using medical records, hospital discharge summaries, death certificates, and autopsy data. Events adjudicated using ICD-10 criteria. |
| **ORIGIN (2012)[25]** | Outcome Reduction with an Initial Glargine Intervention<br><br>NCT00069784 | Unpublished data.<br><br>Data acquired directly from PI. | AF not a prespecified outcome in ORIGIN. PI confirmed AF was reported by individual study sites as part of safety monitoring and was not centrally adjudicated. Participants were followed at routine visits at 2-, 4-, 8-, and 16-wks post-randomization and every 4 mos thereafter, during which safety events were collected. |

| **Low Risk High Dose (LR-HD)** | | | |
|---|---|---|---|
| **RCT-EPA (2023)[26]** | Effects of EPA on Prostate Cancer Cells Proliferation and Quality of Life (RCT-EPA)<br><br>NCT02333435 | Published data.<br><br>Data reported in research article[26]. | AF not a prespecified outcome in RCT-EPA. The study protocol[27] reported that AEs would be collected and monitored at each post-prostatectomy visit using NIH AE criteria (v4.03, June 2010)[28], which include CV/arrhythmic events. In the published article, all AEs were listed, and none were cardiac events. Therefore, AF considered absent by us (0 events). |
| **Mengelberg et al (2022)[29]** | In older adults with mild cognitive impairment, do omega-3 fatty acids compared to a placebo improve cognitive performance?<br><br>ACTRN12614000874617 | Unpublished data.<br><br>Data acquired directly from PIs. | AF was not a prespecified outcome in Mengelberg et al. PI confirmed that any AF events would have been captured under AEs which were monitored throughout the study period, with collection at scheduled assessments (6, 9, and 12 mos). PI confirmed no AF was reported by patients- this was coded as zero events in our analysis. |
| **seAFOod Polyp Prevention (2019)[30, 31]** | The seAFOod (Systematic Evaluation of Aspirin and Fish Oil) polyp prevention trial<br><br>ISRCTN05926847 | Published data.<br><br>Data extracted from research article[30] and NIHR Report[31]. | AF was not a prespecified outcome of the seAFOod Polyp Prevention Trial. However, AEs were a prespecified secondary outcome and were systematically collected according to standard CTIMP pharmacovigilance procedures. AEs were coded using MedDRA system organ classes[6], under which AF falls within the cardiac disorders category. Therefore, while AF was not defined as a specific endpoint, any AF events that occurred were captured as part of routine AE monitoring. Safety analyses were performed in the safety population (all participants who received at least one dose of investigational medicinal product).<br><br>Telephone consultations were conducted at 2, 12, and 38 weeks after randomization to assess AEs, in addition to routine in-person follow-up[32]. PI confirmed that no systematic AF screening (e.g., ECG monitoring) was performed. |

| | | | |
|---|---|---|---|
| **ENrGISE (2019)[33, 34]** | The ENRGISE (ENabling Reduction of Low-Grade Inflammation in SEniors) Study<br><br>NCT02676466 | Published data.<br><br>Data extracted from ClinicalTrials.gov AEs records[34] | AF not a prespecified outcome in ENrGISE. AF events were captured under both SAEs (2 hospitalizations)[34] and non-SAEs (2 events)[34]. AEs were systematically collected from each participant beginning at screening and continuing through study closeout, with quarterly in-person assessments (visits where clinical evaluations, labs, safety reviews, and participant interviews occurred) and allowed participants to report events between visits[33]. |
| **ADCS-DHA (2010)[35, 36]** | Alzheimer's Disease Cooperative Study<br><br>NCT00440050 | Unpublished data.<br><br>Data were extracted from the AEs raw dataset, accessed via formal data application request to ADCS data repository[36] | AF was not a prespecified outcome in ADCS-DHA. PI confirmed AF events were captured under AEs and collected systematically every 3 mos during treatment (follow-up visits at 3, 6, 9, 12, 15, and 18 mos). AF ascertained through multiple routes, including medical records, ECGs, self-report, and other clinical documentation. |
| **MARINA (2011)[†37]** | Vascular effects of eicosapentaenoic acid (EPA) and docosahexaenoic acid (DHA): the MARINA study<br><br>ISRCTN66664610 | Unpublished data.<br><br>Data acquired directly from PIs. | AF not a prespecified outcome of MARINA. PI checked raw dataset (AEs, reasons for non-compliance, dropouts) and confirmed no cases of AF were reported during the study. Although AF was not an outcome, the incidence of AF would have been detected by 24h heart rate monitoring with an Actiheart device (Cambridge Neurotechnology Ltd, Cambridge, UK) that was conducted at baseline, 6- and 12 mos of treatment. PI is confident that no cases of AF were missed. |
| **Lin et al (2022)[†38]** | The Protective Effect of Omega-3 Fatty Acid on Cognitive Function Among Patients with Mild Dementia<br><br>NCT04972643 | Unpublished data.<br><br>Data acquired directly from PIs. | AF not a prespecified outcome of Lin et al. PI confirmed that any AF events would have been captured under AEs reporting. Participants were followed over a 24-month treatment period, with scheduled in-person visits every 6 mos (6, 12, 18, and 24 mos) for AE monitoring. No AF events were reported by participants, and we coded AF incidence as zero in our analysis. |

| | | | |
|---|---|---|---|
| **BRAVE-EPA (2025)[39]** | Brain Amyloid and Vascular Effects of Eicosapentaenoic Acid<br><br>NCT02719327 | Unpublished data.<br><br>Data acquired directly from PIs. | AF not a prespecified outcome in BRAVE-EPA. PI confirmed AF recorded under AEs, based on patient self-report. Participants were assessed for AEs at mos 1, 3, 6, 9, 12, 15, and 18. |
| **EPOCH (2018)[40, 41]** | The Older People, Omega-3, and Cognitive Health (EPOCH) trial<br><br>ACTRN12607000278437 | Unpublished data.<br><br>Data acquired directly from PI. | AF not a prespecified outcome in EPOCH. PI confirmed AF recorded under AEs, based on patient self-report. According to trial protocol[41] adverse effects suspected as being related to supplement use recorded by research officer at visits every 3 mos during the 18-month intervention period. Events reviewed by trial physician as needed. |
| **DREAM (2019)[42]** | The Dry Eye Assessment and Management<br><br>NCT02128763 | Published data.<br><br>Data extracted from supplemental table[42]. | AF not a prespecified outcome in DREAM. AF (2 events) and atrial flutter (1 event) was self-reported and recorded as part of SAEs reporting categorized under cardiac disorders. Coordinators systematically asked participants about AEs during each in-person visit (at 3, 6, and 12 mos) and by telephone at 9 mos, coding all events using the Medical Dictionary for Regulatory Activities (MedDRA, version 10.0)[43] |
| **CAPFISH-3 (2024)[44]** | Low-Fat Diet and Fish Oil in Men on Active Surveillance for Prostate Cancer<br><br>NCT02176902 | Unpublished data.<br><br>Data acquired directly from PIs. | AF not a prespecified outcome of CAPFISH-3. AF was self-reported and recorded as part of AE reporting during in-person and phone visits monthly for the omega-3 group and in-person visits at 6 and 12 mos for the control group. Incidence of AEs graded according to according to National Cancer Institute Common Toxicity Criteria version 4.0[45]. |

| | | | |
|---|---|---|---|
| **Low Risk Low Dose (LR-LD)** | | | |
| **Shinto et al (2014)[46, 47]** | Lipoic Acid and Omega-3 Fatty Acids for Alzheimer's Disease<br><br>NCT01058941 | Published data.<br><br>Data extracted from ClinicalTrials.gov AEs records[46] | AF not a prespecified outcome in Shinto et al (2014). Events were collected by systematic assessment and captured under AEs which were monitored monthly from day 0 through 18 mos, during clinic visits and by phone. Safety monitoring included participant and study partner reports, laboratory tests, vital signs, and physical examinations. |
| **VITAL Rhythm (2021)[‡48]** | VITamin D and Omega-3 Trial – Rhythm Study<br><br>NCT01169259 | Published data.<br><br>Data extracted from supplemental table[48] | AF reported as a prespecified primary endpoint of VITAL-Rhythm ancillary study. AF or flutter events were identified by participant self-report on annual questionnaires and by claims data linkage (via CMS / insurance claims using ICD-9/ICD-10 codes). For all potential AF or flutter cases thus identified, the study requested permission to obtain relevant medical records. An endpoint adjudication committee (cardiologists / expert reviewers) then confirmed events based on ECG evidence or physician documentation of AF or atrial flutter. If participants did not return questionnaires each year, they were presumed not to have developed AF unless subsequently identified via claims or records. |
| **CANN (2023)[49]** | Cognitive Ageing Nutrition and Neurogenesis<br><br>NCT02525198 | Unpublished data.<br><br>Data acquired directly from PIs. | AF not a prespecified outcome of CANN. PI confirmed that if patients experienced AF, it would have been captured under AEs which were recorded at in-person visits at 3- and 12- mos. As no AF was reported by patients, we coded this as zero events in our analysis. |

| | | | |
|---|---|---|---|
| **AREDS2 (2013)**[50, 51] | Age-Related Eye Disease Study 2<br><br>NCT00345176 | Unpublished data.<br><br>Data were extracted from the AEs raw dataset[51], obtained through internal collaboration with the trial investigators. | AF was a predefined secondary safety endpoint in AREDS2, captured under SAEs. PI confirmed CV events, including AF, were initially self-reported by participants, with medical record review when available. Formal adjudication of CVD events was performed by a medical committee of cardiologists, internists, and neurologists. |
| **MARINA (2011)**[†37] | Vascular effects of eicosapentaenoic acid (EPA) and docosahexaenoic acid (DHA): the MARINA study<br><br>ISRCTN66664610 | Unpublished data.<br><br>Data acquired directly from PIs. | Same information as detailed in LR-HD group[†] |
| **Lin et al (2022)**[†38] | The Protective Effect of Omega-3 Fatty Acid on Cognitive Function Among Patients with Mild Dementia<br><br>NCT04972643 | Unpublished data.<br><br>Data acquired directly from PIs. | Same information as detailed in LR-HD group[†] |
| **DO-HEALTH (2025)**[52] | VitaminD3-Omega3-Home Exercise-HeALTHy Ageing and Longevity Trial<br><br>NCT01745263 | Unpublished data.<br><br>Data acquired directly from PI. | AF was a predefined safety endpoint in DO-HEALTH. AF events were captured through systematic AE monitoring, which included structured phone interviews every 3 mos throughout the 3-yr follow-up, as well as in-person clinic visits at baseline and at 12, 24, and 36 mos. Reported events were classified using ICD-10 (2010 v) terminology within the safety monitoring system. |

Abbreviations: AF: atrial fibrillation; AEs: adverse events; CRF: case report form; CTIMP: clinical trial of an investigational medicinal product; CV: cardiovascular; DHA: docosahexaenoic acid; DSMB: Data and Safety Monitoring Board; ECGs: electrocardiograms; EPA: eicosapentaenoic acid; HF: heart failure; ICD: implantable cardioverter-defibrillator; mos: months; NIH: National Institutes of Health; N: number of participants randomized; PIs: principal investigators; RCT: randomized controlled trial; SAEs: serious adverse events; wks: weeks; yr: year.

[†]MARINA and Lin et al., investigated multiple doses of omega-3 in low-risk participants. As a result, their low-dose arms were included in the LR-LD category, and their high-dose arms were included in the LR-HD category in the stratified analyses.

[‡]VITAL-rhythm is an ancillary trial of the VITAL trial (NCT01169259)- our meta-analysis was performed using VITAL-rhythm data.
