## Supplemental Table 2 for "Effects of Omega-3 Fatty Acid Treatment on Risk for Atrial Fibrillation: An Updated Meta-Analysis of 34 Trials including 114,326 Individuals"

**Supplemental Table 2.** RCTs meeting eligibility criteria but excluded from the meta-analysis due to principal investigators (PIs)/corresponding authors were unable to provide data or did not respond to repeated requests (after 3x attempts at follow-up).

| Trial/Authors<br><br>(Year) | Trial Full-name<br><br>Trial Registration No. (if applicable) | Intervention<br><br>Study Duration (Months) | Sample Size (N)<br><br>Study Population | PIs Response |
|---|---|---|---|---|
| **Responded- No relevant data collected** | | | | |
| **FISH (2012)[1]** | Fish oil Inhibition of Stenosis in Hemodialysis grafts study<br><br>ISRCTN15838383<br>Canada and United States | MEG-3® 4000mg/d (EPA 1600mg/d + 800mg/d DHA)<br>Placebo (corn oil)<br><br>12 months | N=232<br><br>Chronic hemodialysis patients who needed new graft access. | PI confirmed that AF was not a pre-specified outcome of FISH; no relevant data collected. |
| **Nosaka et al (2017)[2]** | Clinical effect of early loading of eicosapentaenoic acid after percuta-neous coronary intervention in patients with acute coronary syndrome<br><br>UMIN000016723<br>Japan | Vascepa 1800mg/d (1800mg/d EPA +2mg/d Pitavas-tatin)<br>Control (2mg/d Pitavastatin)<br><br>12 months. | N=241<br><br>Patients w/ acute coronary syndrome patients treated w/ successful percutaneous coronary intervention within 24 hrs. | PI confirmed they do not have data on incidence AF. |
| **WELCOME (2014)[3]** | Wessex Evaluation of fatty Liver and Cardiovascular markers in NAFLD with Omacor thErapy<br><br>NCT00760513<br>United Kingdom | Omacor 4000mg/d (1840mg/d EPA + 1520mg/d DHA)<br>Placebo (olive oil)<br><br>18 months. | N=103<br><br>Patients w/non-alcoholic fatty liver disease. | PI confirmed that AF was not a pre-specified outcome of WELCOME; no data collected. |
| **Baleztena et al (2018)[4]** | Dietary Supplement for the Prevention of Cognitive Decline in a Very Elderly Population.<br><br>NCT01817101<br>Spain | Omega-3 1050 mg/d (120mg/d EPA + 750mg/d DHA)<br>Placebo (gelatin).<br><br>12 months | N=99<br><br>≥75-year-old nursing home resident's w/o CI or with mild cognitive impairment (MCI). | PI confirmed AF was not recorded and was not a pre-specified outcome; access to EMRs for AF assessment was not possible due to lack of consent and participant availability. |

| | | | | |
|---|---|---|---|---|
| **SOFA (2006)**[5] | The Study on Omega-3 Fatty acids and ventricular Arrhythmia<br><br>NCT00110838<br>Europe | Omega-3 2000mg/d (EPA 450mg/d + DHA 350mg/d)<br>Placebo (inert oil unspecified)<br><br>12 months | N=546<br><br>Patients w/implantable cardioverter defibrillators and prior documented malignant ventricular tachycardia (VT) or ventricular fibrillation (VF). | PI confirmed all arrhythmias captured under AEs were ventricular (ICD-detected); AF could not be isolated from VF or VF. Therefore, no relevant data collected in SOFA. |
| **OPAL (2010)**[6] | The Older People And n-3 Long-chain polyunsaturated fatty acids study<br><br>United Kingdom<br>ISRCTN72331636 | Omega-3 1000mg/d (EPA 200mg/d + DHA 500mg/d)<br>Placebo (olive oil)<br><br>24 months | N=867<br><br>Cognitively healthy adults aged 70-79 yrs. | PI confirmed that AF was not a pre-specified primary or secondary outcome of OPAL and that no relevant data are available for inclusion. Follow-up regarding AEs data also confirmed that no AF-related information was collected. |
| **The Beyond Ageing Project (2017)**[7] | The Beyond Ageing Project: A selective prevention trial using novel pharmacotherapies in an older age cohort at risk for depression<br><br>ACTRN12610000032055<br>Australia | Omega-3 2000mg/d (1200mg/d EPA + 800mg/d DHA) or 50mg/d Sertraline hydrochloride<br>Placebo (paraffin oil).<br><br>12 months | N=450<br><br>A community-based sample subjects w/previously reported sub-threshold depressive symptoms, aged 60-74 yrs. | PI confirmed AF was not a pre-specified outcome of Beyond Ageing Project; no relevant data collected. |
| **NICE (2024)**[8] | Nutrition Interventions for Cognitive Enhancement (NICE)<br><br>NCT03841539<br>United States | Omega-3 2000mg/d (ratio EPA/DHA unspecified) combined w/ dietary counselling (Mediterra-nean or Low-fat).<br>Placebo (unspecified)<br><br>12 months | N=209<br><br>Cognitively normal older adults aged ≥65 yrs. | PI confirmed that AF was not a pre-specified outcome in NICE and that no relevant AF data were collected. |
| **IRIS-FAI (2024)**[9] | Effect of Icosapent ethyl on inflammation in the vessel wall assessed by the Fat Attenuation Index Score<br><br>ISRCTN15140257<br>United Kingdom | Vazkepa® (icosapent ethyl) x2 capsules (1920mg/d EPA) combined w/ standard care<br>Control (standard care only)<br><br>12 months | N=100 (target)<br><br>High-risk adults w/ established cardiovascular disease and elevated triglyceride levels | The PI was unable to assist. The study was terminated in October 2025 owing to recruitment challenges. |

| **Responded-Relevant data collected but not shareable** | | | | |
|---|---|---|---|---|
| **PISCES (2025)[10]** | Protection against Incidences of Serious Cardiovascular Events Study with daily fish oil supplementation in dialysis patients<br><br>ISRCTN00691795<br>Canada and Australia | Omega-3 4000mg/d<br>(EPA 1600mg/d + DHA 800mg/d)<br>Placebo (corn oil)<br><br>42 months | N=1100<br><br>Patients w/ end-stage kidney disease who requires chronic hemodialysis 3 or 4 times/wk. | PI confirmed that arrhythmia data including atrial and ventricular arrhythmias were collected in PISCES; however, these data are not currently shareable. The team plans to present the arrhythmia results in March 2026. |
| **PreventE4 (2023)[11]** | The Brain DHA Delivery Trial<br><br>NCT03613844<br>United States | Omega-3 2000mg/d<br>(DHA 2000mg/d)<br>Placebo (corn and soy oil)<br><br>24 months | N=365<br><br>Cognitively unimpaired individuals aged 55-80 yrs, with at least one CV or dementia risk factor (obesity, hypertension, hyperlipidemia, or physical inactivity). | PI confirmed that AF was recorded as an AE but could not provide the data until the main trial results are published in a peer-reviewed journal. |
| **NUTRIMEMO (2015)[12,13]** | Effect of Lipidic Nutrients on Memory and Well Being in Healthy Aging Adults<br><br>NCT02626247<br>France | Omega-3 combined w/ Vitamin A x3 capsules/d<br>(EPA+DHA 900mg/d)<br>Placebo (unspecified)<br><br>12 months | N=340<br><br>Healthy elderly subjects independently living at home aged 60-70 yrs. | PI confirmed that AF was recorded as an AE but could not provide the data until the main trial results are published in a peer-reviewed journal. |

| | | | | |
|---|---|---|---|---|
| **PUFA (2024)**[14] | Prevention of Cerebral Small Vessel Disease and Inflammation in Aging<br><br>NCT01953705<br>United States | Omega-3 1650mg/d<br>(EPA 975mg/d + DHA 650mg/d)<br>Placebo (soybean oil)<br><br>36 months | N=102<br><br>Participants w/o dementia but with mild cognitive impairment aged 75 yrs and older. | PI recalled 21 abnormal ECGs at 12 months (≈10 in omega-3 group, ≈11 in placebo), with no between-group differences. Reasons for abnormalities unclear; confirmation would require reading ECGs. The PI initially agreed to revisit the ECGs to verify whether any represented AF; however, no further responses were received despite follow-up attempts. |
| **Responded-AF data status unclear** | | | | |
| **DOIT (2010)**[15] | Diet and Omega-3 Intervention Trial (DOIT) on atherosclerosis<br><br>Norway | Pikasol® 2400mg/d<br>(EPA 840mg/d + DHA 480mg/d) combined w/ or w/o dietary counselling (Mediterranean)<br>Control (dietary counselling)<br>Placebo (corn oil).<br><br>36 months. | N= 563<br><br>Elderly men aged 65-75 yrs w/ longstanding dyslipidemia or hypertension. | Co-investigator confirmed that AF was not a prespecified outcome of DOIT and indicated they do not have access to the raw dataset to review AEs. The PI may hold the dataset, but multiple contact attempts by us received no response. |
| **MAPT (2019)**[16] | Omega-3 Fatty Acids and/or Multi-domain Intervention in the Prevention of Age-related Cognitive Decline<br><br>NCT00672685<br>France | Omega-3 1025mg/d<br>(800mg/d DHA + 225mg/d EPA) combined w/ or w/o multi-domain intervention (nutrition, physical exercise, cognitive stimulation, social activities).<br>Placebo (unspecified)<br><br>36 months | N=1680<br><br>Community-dwelling participants aged ≥70 yrs non-demented, but w/ memory complaint and limitations in one instrumental activity of daily living, or slow gait speed. | PI confirmed that AF was not a pre-specified outcome of MAPT and that AF cases may be identifiable within AE records, however this analysis has not been conducted. The PI did not respond to follow-up requests asking whether AE data could be re-viewed for AF events. |

| | | | | |
|---|---|---|---|---|
| **NAT2 (2013)**[17] | Nutritional AMD Treatment 2 Study<br><br>ISRCTN98246501<br>France | Omega-3 840mg/d<br>(DHA 840mg/day)<br>Placebo (olive oil)<br><br>36 months | N=263<br><br>Patients aged 55-85 yrs w/early lesions of age-related maculopathy (at risk of age-related macular degeneration). | PI was unable to assist and provided no further information. It remains unclear whether AF-related data were collected or exist within the study records. |
| **JELIS (2007)**[18] | Japan EPA Lipid Intervention Study<br><br>NCT00231738<br>Japan | Vascepa® 1800mg/d combined w/ 10mg/d Pravastatin or 5mg/d Simvastatin<br>(EPA 1800mg/d)<br>Control (Statin therapy alone)<br><br>55 months (mean follow-up) | N= 18,645<br><br>Patients' w/ Hypercholesterolemia undergoing statin therapy. | Co-investigators confirmed that AF was not a pre-specified outcome of JELIS and do not have access to the raw dataset to review AEs. |
| **EPE-A (2014)**[19] | Double-Blind, Placebo-Controlled Study of Two Doses of EPA-E in Patients With Non-Alcoholic Steatohepatitis<br><br>NCT01154985<br>United States | Omega-3 1800mg/d or 2700mg/d<br>(EPA 1800mg/d or 2700mg/d)<br>Placebo (unspecified)<br><br>12 months | N=243<br><br>Patients w/nonalcoholic steatohepatitis and/or non-alcoholic fatty liver disease. | PI confirmed that AF was not a pre-specified outcome of EPE-A. PI does not have access to EPE-A raw dataset; data are held exclusively by the study sponsor. |
| **Re-MIND (2022)**[20] | Memory Intervention with Nutrition for Dementia<br><br>ISRCTN11892249<br>Ireland | Omega-3 1000mg/d<br>(DHA 500mg/d + EPA 150mg/d)<br>Placebo (sunflower oil)<br><br>24 months | N=120<br><br>Patients w/ mild-moderate Alzheimer Disease. | Discussion with PI planned to address IRB ethical approval for secondary data use; follow-up was made, but received no response. |
| **No response** | | | | |
| **OMEGA (2013)**[21] | Effect of Omega 3-Fatty Acids on the Reduction of Sudden Cardiac Death After Myocardial Infarction<br><br>NCT00251134<br>Germany | Omega-3 1000mg/d<br>(EPA 460mg/d + DHA 380mg/d)<br>Placebo (olive oil)<br><br>12 months | N=3851<br><br>Patients who survived acute myocardial infarction. | New-onset AF data found in systematic review[22] which required further clarification from PIs who did not respond to our request. |

| | | | | |
|---|---|---|---|---|
| **SHOT (1996)[23]** | Scandinavian Heart Omega-3 Trial<br><br>Denmark | Omacor® 4000mg/d combined w/ 15mg/d Warfarin or 300mg/d Aspirin<br>(1840mg/d EPA + 1520mg/d DHA)<br>Placebo (margarine)<br><br>12 months | N=610<br><br>Patients admitted for coronary bypass grafting surgery. | No response. |
| **Tande et al (2016)[24]** | Clinical safety evaluation of marine oil derived from Calanus finmarchicus<br><br>Norway | Calanus oil 2000mg/d<br>(EPA/DHA dosage unspecified)<br>Placebo (unspecified)<br><br>12 months | N=127<br><br>Healthy subjects with BMI 25-35 kg/m$^2$ | No response. |
| **AQUAMARINE EPA/DHA (2024)[25]** | The Attempts at Plaque Vulnerability Quantification with Magnetic Resonance Imaging Using Non contrast T1-weighted Technic EPA/DHA study<br><br>Japan<br>UMIN 000015316 | Lotriga® 2000mg/d or 4000mg/d<br>(EPA 930mg/d + DHA 750mg/d or EPA 1860mg/d + 1500mg/d)<br>Control (standard statin therapy)<br><br>12 months | N=84<br><br>Patients' w/ CAD receiving statin therapy. | No response. |
| **Duan et al (2025)[26]** | Effects of medium chain triglycerides combined with DHA on ameliorating cognitive decline by brain energy metabolism in the elderly with mild cognitive impairment<br><br>ChiCTR2200059641<br>China | Algae oil 2000mg/d<br>(DHA 900-1100mg/d)<br>Placebo (corn oil)<br><br>24 months | N=240<br><br>Elderly individuals with mild cognitive impairment. | No response. |
| **SCIMO (2001)[27]** | Study on Prevention of Coronary Atherosclerosis with Marine Omega-3 Fatty Acids<br><br>Netherlands | Omega-3 6000mg/d (1650mg/d EPA + DHA) for the first 3 months then for the following 21 months 3000mg/d (1500mg/d EPA + DHA)<br>Placebo (composition reflecting average European diet).<br><br>24 months | N=223<br><br>Patients w/angiographically proven CAD. | No response. |

| | | | | |
|---|---|---|---|---|
| **PNMD (2025)**[28] | The Precision Nutritional Management for Diabetes trial<br><br>NCT03708887<br>China | Omega-3 3000mg/d or 1500mg/d (EPA 2160mg/d + DHA 840mg/d or EPA 1080mg/d + DHA 420mg/d)<br>Placebo (olive oil)<br><br>12 months | N=415<br><br>Patients w/ type-2 diabetes or obesity. | No response. |
| **PAOXRED (2022)**[29] | Protección AntiOXidante en la REtinopatía Diabética (Antioxidant Protection in Diabetic Retinopathy)<br><br>Spain | Brudyretina 1500mg/d (EPA 127mg/d + DHA 1050mg/d)<br>Placebo (olive oil)<br><br>24 months | N=170<br><br>Patients w/non-proliferative diabetic retinopathy. | No response. |
| **Garcia Layana et al (2021)**[30] | Study of Nutritional Supplementation in Patients With Unilateral Wet AMD<br><br>Spain<br>NCT04756310 | Retilut® capsules/d (DHA 800mg/d)<br>Control (Theavit® nutritional supplement w/o DHA)<br><br>12 months | N=109<br><br>Patients w/diagnosed unilateral exudative age-related macular degeneration. | No response. |
| **Ahn et al (2016)**[31] | Effect of n-3 Polyunsaturated Fatty Acids on Regression of Coronary Atherosclerosis in Statin Treated Patients Undergoing Percutaneous Coronary Intervention<br><br>Korea | Omega-3 (3000mg/d) (EPA 1395mg/d + DHA 1125mg/d)<br>Placebo (unspecified)<br><br>12 months. | N=74<br><br>Patients undergoing percutaneous coronary intervention w/ stent implantation and undergoing statin therapy. | No response. |
| **Raitt et al (2005)**[32] | Fish Oil Supplementation and Risk of Ventricular Tachycardia and Ventricular Fibrillation in Patients With Implantable Defibrillators<br>A Randomized Controlled Trial<br><br>United States | Omega-3 1800mg/d (EPA 756mg/d + DHA 540mg/d)<br>Placebo (olive oil)<br><br>24 months | N=200<br><br>Patients with implantable cardioverter defibrillators and recent episode of sustained VT or VF. | A total of 47 AF events were identified through ICD electrogram review; however, it is unclear whether these represented new-onset or pre-existing AF, and the allocation of events to the omega-3 or placebo groups could not be determined. Attempts to obtain clarification from the PI received no response. |

| | | | | |
|---|---|---|---|---|
| **Brox et al (2001)**[33] | A Long-Term Seal- and Cod-Liver-Oil Supplementation in Hypercholesterolemic Subjects<br><br>Norway | Cod-liver oil 15ml/d<br>(EPA 1500mg/d + DHA 1800mg/d)<br>or Seal oil 15ml/d<br>(EPA 1100mg/d + DHA 1500mg/d)<br>Control (nothing given)<br><br>14 months | N=120<br><br>Subjects with moderate hypercholesterolemia. | No response. |
| **Sandhu et al (2016)**[34] | Nutritional Supplements and Hormonal Manipulations for Breast Cancer Prevention<br><br>NCT00723398<br>United States | Lovaza® 4000mg/d alone or combined w/ 30mg Raloxifene<br>(EPA 1860mg/d + DHA 1500mg/d)<br>or 30mg Raloxifene alone<br>or 60mg Raloxifene alone<br>Control (nothing given)<br><br>24 months | N=266<br><br>Healthy postmenopausal women at increased risk of breast cancer based on high breast density detected on their routine screening mammograms. | No response. |
| **Puri et al (2005)**[35] | A multicentre, multinational, double blind, randomised, parallel group, placebo-controlled study of ethyl-eicosapentaenoate (EPA) in patients with Huntington's disease (HD)<br><br>ISRCTN79170611<br>United Kingdom | Omega-3 2000mg/d<br>(EPA 2000mg/d)<br>Placebo (unspecified)<br><br>12 months | N=135<br><br>Patients w/ Huntington disease. | No response. |
| **Sabour et al (2015)**[36] | Omega 3 in Intervention Spinal Cord Injured People<br><br>Iran<br>NCT01311375 | MorDHA® x2 capsules/d<br>(EPA 130mg/d + DHA 870mg/d)<br>Placebo (unspecified)<br><br>14 months. | N=110<br><br>Chronic traumatic spinal cord-injured patients. | No response. |
| **Olendzki et al (2011)**[37] | Treatment of Rheumatoid Arthritis with Marine and Botanical Oils: Influence on Serum Lipids<br><br>United States | Omega-3 3500mg/d combined w/ 1800mg/d borage oil (GLA)<br>(2100mg/d EPA + 1400mg/d DHA)<br>Placebo (sunflower oil)<br><br>18 months | N=146<br><br>Rheumatoid arthritis patients w/ active joint inflammation' | No response. |

| | | | | |
|---|---|---|---|---|
| **OPACH (2006)**[38] | Omacor in Prevention of Cardiovascular Events in Patients Undergoing Chronic Hemodialysis<br><br>Denmark<br>NCT00257283 | Omacor® 2000mg/d<br>(EPA 920mg/d + DHA 760mg/d)<br>Placebo (olive oil)<br><br>24 months | N=206<br><br>Patients w/ documented CVD, undergoing chronic hemodialysis for at least 6 mos. | No response. |
| **HOST (2016)**[39] | The Impact of Omega Three Fatty Acids on Vascular Function in HIV<br><br>NCT01041521<br>United States | Lovaza® 4000mg/d<br>(EPA 1860mg/d + DHA 1500mg/d)<br>Placebo (sugar pill)<br><br>24 months | N=129<br><br>HIV-infected individuals with elevated levels of triglycerides (≥150 mg/dl). | No response. |
| **Li et al (2024)**[40] | Cognitive Benefits of Folic Acid, Docosahexaenoic Acid, and a Combination of Both Nutrients in Mild Cognitive Impairment: Possible Alterations through Mitochondrial Function and DNA Damage<br><br>China | Omega-3 800mg/d combined w/ or w/o 800μg/d folic acid<br>(DHA 800mg/d)<br>Placebo (unspecified)<br><br>12 months. | N=280<br><br>MCI participants aged 60 yrs and older. | No response. |
| **Lafuente et al (2017)**[41] | Three-Year Outcomes in a Randomized Single-Blind Controlled Trial of Intravitreal Ranibizumab and Oral Supplementation with Docosahexaenoic Acid and Antioxidants For Diabetic Macular Edema<br><br>EudraCT 2015-001082-74<br>Spain | Brudyretina® 1500mg/d combined w/ 0.5mg/d Ranibizumab<br>(EPA 127mg/d + DHA1050mg/day)<br>Control (0.5mg/d Ranibizumab)<br><br>36 months | N=55<br><br>Patients w/ type 2 diabetes who presented decreased vision due to central-involved diabetic macular edema. | No response. |

| | | | | |
|---|---|---|---|---|
| **NutriStroke (2009)**[42] | The Role of Nutrition in the Rehabilitation of Patients With Eating Disorders After a Vascular Stroke<br><br>Italy | Omega-3 500mg/d combined w/ or w/o antioxidants<br>(EPA 250mg/d + DHA 250mg/d)<br>Placebo (identical supplement but contained no antioxidants or omega-3)<br><br>12 months. | N=72<br><br>Stroke patients admitted to a rehabilitation hospital for sequelae of first-ever ischemic stroke. | No response. |
| **CARES (2022)**[43] | Cognitive impAiRmEnt Study: Changes in brain function among individuals with a mild memory impairment<br><br>ISRCTN10431469<br>Ireland | Omega-3 1000mg/d combined w/ carotenoids (lutein, zeaxanthin, and meso-zeaxanthin)<br>(DHA 430mg/d + EPA 90mg/d)<br>Placebo (sunflower oil)<br><br>24 months | N=60<br><br>Individuals w/ mild cognitive impairment aged ≥65 yrs. | No response. |
| **Blok et al (1997)**[44] | Pro- and anti-inflammatory cytokines in healthy volunteers fed various doses of fish oil for 1 year<br><br>The Netherlands | Omega-3 3000mg/d<br>(EPA+DHA 1060mg/d) or<br>Omega-3 6000mg/d<br>(EPA+DHA 2130mg/d) or<br>Omega-3 9000mg/d<br>(EPA+DHA 3190mg/d)<br>Placebo (identical supplement w/o omega-3).<br><br>12 months | N=58<br><br>Healthy monks (mean age 56 yrs). | No response. |
| **SALVAGE (2024)**[45] | IcoSApent ethyL to Slow Down Aortic VAlve Stenosis proGrEs-sion (SALVAGE)<br><br>NCT06466278<br>The Netherlands | Vazkepa® x4 capsules/d<br>(3840mg/d EPA)<br>Placebo (unspecified)<br><br>24 months | N=110 (estimated)<br><br>Patients w/ mild to moderate to aortic valve stenosis aged ≥50 yrs. | No response. |

| | | | | |
|---|---|---|---|---|
| **TotAL (2018)**[46] | Testosterone-Omega Three (DHA) - Amyloid Lowering (TotAL) study<br><br>Australia<br>ACTRN12618000761268 | Omega-3 x4 capsules/d (1720mg DHA) combined w/ or w/o intramuscular testosterone injections every ~12 weeks<br>Placebo (corn oil)<br><br>14-18 months approx. | N=200<br><br>Males aged between 60-80 yrs w/ subjective memory complaints, and amyloid-positive PET scans. | No response. |

Abbreviations: AF: atrial fibrillation; AEs: adverse events; CAD: coronary artery disease; CV: cardiovascular; CVD: cardiovascular disease; DHA: docosahexaenoic acid; ECGs: electrocardiograms; EPA: eicosapentaenoic acid; HF: heart failure; HIV: human immunodeficiency virus; ICD: implantable cardioverter-defibrillator; IRB: institutional review board; MCI: mild cognitive impairment; mg/d: milligrams per day; mos: months; N: number of participants randomized; PET: positron emission tomography; PIs: principal investigators; RCT: randomized controlled trial; SAEs: serious adverse events; VT: ventricular tachycardia; VF: ventricular fibrillation; wks: weeks; yr: year.

39- Gretchen Volpe, Sally Skinner, Jul Gerrior-Schofield, Fang Fang Zhang, Alice Tang, Jeffrey Kuvin, Alexandra Mangili, Christine Wanke, A Randomized Controlled Trial of Omega-3 Fatty Acids in Human Immunodeficiency Virus (HIV): Long-Term Effects on Lipids and Vascular Function, Open Forum Infectious Diseases, Volume 3, Issue suppl_1, December 2016, 951,

40- Li M, Li T, Yang T, Huang L, Zhao J, Liu H, Chen Y, Li W, Zhu Y, Ma F, Yan J, Huang G. Cognitive Benefits of Folic Acid, Docosahexaenoic Acid, and a Combination of Both Nutrients in Mild Cognitive Impairment: Possible Alterations through Mitochondrial Function and DNA Damage. Gerontology. 2024;70(9):940-949. doi: 10.1159/000540021. Epub 2024 Jul 17. PMID: 38952108.

41- Lafuente, M., Ortín, L., Argente, M., Guindo, J.L., López-Bernal, M.D., López-Román, F.J., Domingo, J.C. and Lajara, J., 2019. Three-year outcomes in a randomized single-blind controlled trial of intravitreal ranibizumab and oral supplementation with docosahexaenoic acid and antioxidants for diabetic macular edema. Retina, 39(6), pp.1083-1090.

42- Garbagnati F, Cairella G, De Martino A, Multari M, Scognamiglio U, Venturiero V, Paolucci S. Is antioxidant and n-3 supplementation able to improve functional status in poststroke patients? Results from the Nutristroke Trial. Cerebrovasc Dis. 2009;27(4):375-83. doi: 10.1159/000207441. Epub 2009 Mar 10. PMID: 19276620.

43- Power, R., Nolan, J.M., Prado-Cabrero, A., Roche, W., Coen, R., Power, T. and Mulcahy, R., 2022. Omega-3 fatty acid, carotenoid and vitamin E supplementation improves working memory in older adults: A randomised clinical trial. Clinical Nutrition, 41(2), pp.405-414.

44- Blok WL, Deslypere JP, Demacker PN, van der Ven-Jongekrijg J, Hectors MP, van der Meer JW, Katan MB. Pro- and anti-inflammatory cytokines in healthy volunteers fed various doses of fish oil for 1 year. Eur J Clin Invest. 1997 Dec;27(12):1003-8. doi: 10.1046/j.1365-2362.1997.2240775.x. PMID: 9466128.

45- IcoSApent ethyL to Slow Down Aortic VAlve Stenosis proGrEssion (SALVAGE) (2023) ClinicalTrials.gov NCT06466278. Available at: https://clinicaltrials.gov/ct2/show/NCT06466278 (Accessed: 06 October 2025).

46- Testosterone-Omega Three (DHA) — Amyloid Lowering (ToTAL) Study (2018) Australian New Zealand Clinical Trials Registry ACTRN12618000761268. Available at: https://www.anzctr.org.au/Trial/Registration/TrialReview.aspx?id=373908 (Accessed: 06 August 2025).
