## Supplemental Table 3 for "Effects of Omega-3 Fatty Acid Treatment on Risk for Atrial Fibrillation: An Updated Meta-Analysis of 34 Trials including 114,326 Individuals"

**Supplemental Table 3.** Characteristics of the 34 RCTs included in the meta-analysis stratified by cardiovascular risk and omega-3 dosage (high dose >1500 mg E+D/d).

| Trial/ Authors (Year) | Country | N | Study Population | Mean Age<br>Males (%) | Omega-3 Formulation | Intervention and Comparator | M | Ratio (EPA: DHA) |
|---|---|---|---|---|---|---|---|---|
| **High Risk High Dose (HR-HD)** | | | | | | | | |
| **Yokote et al (2020)[1]** | Japan | 383 | Patients' w/ hyperlipidemia accompanied by hypertriglyceridemia. | 56.9<br>78% | O3CA | Epanova® 2000mg/d (EPA 1100mg/d + DHA 400mg/d) Epanova® 4000mg/d (EPA 2200mg/d + DHA 800mg/d<br>Placebo (corn oil) | 12 | 2.75:1 |
| **REDUCE-IT (2023)[2,3]** | Multi-national | 8,179 | Statin-treated patients w/ hypertriglyceridemia, and high risk of/or established ASCVD and/or TD1/2. | 64<br>71% | IPE | Vascepa® 4000 mg/day (EPA 4000mg/d)<br>Placebo (mineral oil) | 58.8 | NA |
| **STRENGTH (2020)[4]** | Multi-national | 13,078 | Statin-treated patients w/ hypertriglyceridemia, low HDL, and high risk of/or established ASCVD and/or TD1/2. | 62.5<br>65% | O3CA | Epanova® 4000mg/d (EPA 2200mg/d + DHA 800mg/d)<br>Placebo (corn oil) | 42 | 2.75:1 |
| **FAAT (2005)[5]** | USA | 402 | Patients' w/ implantable cardioverter/defibrillators at high risk of fatal ventricular arrhythmias. | 65<br>85% | O3AEE | Omacor® 4000mg/d (EPA+DHA 2600 mg/d)<br>Placebo (olive oil) | 12 | 1.24:1 |

| | | | | | | | | |
|---|---|---|---|---|---|---|---|---|
| **PEACH (2018)[6]** | Japan | 157 | Patients' w/ hypercholesterolemia undergoing statin therapy. | 67<br><br>53% | EE | Omega-3 1800mg/d combined w/ Pitavastatin 2mg/d<br>(EPA 1800mg/d)<br><br>Control (Pitavastatin 2mg/d or 4mg/d) | 12 | NA |
| **OMEMI (2023)[7]** | Norway | 1,027 | Elderly patients (70-82 yrs) w/recent MI 2-8 wks prior. | 74<br><br>71% | rTG | Omega-3 1800mg/d<br>(EPA 930mg/d + DHA 660mg/d)<br><br>Placebo (corn oil) | 24 | 1.4:1 |
| **EVAPORATE (2020)[8]** | USA | 80 | Patients w/ coronary atherosclerosis under-going statin therapy, and have persistently elevated triglyceride levels. | 57.4<br><br>54% | IPE | Vascepa® 4000 mg/day<br>(EPA 4000mg/d)<br><br>Placebo (mineral oil). | 18 | NA |
| **HEARTS (2025)[9]** | USA | 285 | Patients' w/stable CAD on statin therapy. | 63<br><br>85% | O3AEE | Lovaza® 3360mg/d<br>(EPA 1860mg/d +DHA 1500mg/d)<br><br>Control (standard care) | 30 | 1.24:1 |
| **SAREPITASO (2018)[10]** | Japan | 76 | Patients w/ T2DM and non-diabetic controls. | 74<br><br>47% | EE | Omega-3 1800mg/d<br>(EPA 1800mg/d combined w/ Pitavastatin 2mg/d)<br><br>Control (Sarpogrelate 300mg/d + Pitavastatin 2mg/d | 12 | NA |
| **Derosa et al (2016)[11]** | Italy | 281 | Overweight/obese patients w/ impaired fasting glucose or impaired glucose tolerance. | 53.5<br><br>50% | EE | Omega-3 3000mg/d combined w/ diet and physical activity.<br>(EPA+DHA dose unspecified)<br><br>Placebo (sucrose, mannitol and mineral salts) | 18 | 0.9:1.5 |
| **OFAMI (2001)[12]** | Norway | 300 | Patients w/ existing CVD following MI (recruited 4-8 days after confirmed MI). | 64<br><br>74% | O3AEE | Omacor® 4000mg/d<br>(~3400-3500 mg/d EPA + DHA)<br><br>Placebo (corn oil) | 18 | 1.24:1 |

| | | | | | | | | |
|---|---|---|---|---|---|---|---|---|
| **RESPECT-EPA (2024)**[13] | Japan | 2,506 | Patients w/ stable CAD and a low EPA/AA ratio. | 68<br><br>80% | IPE | Vascepa® 1800mg/d combined w/ standard statin therapy<br>(EPA 1800mg/d)<br><br>Control (standard statin therapy only) | 60 | NA |
| **High Risk Low Dose (HR-LD)** | | | | | | | | |
| **GISSI-HF (2013)**[14] | Italy | 5,835 | Patients w/heart failure irrespective of cause and left ventricular ejection fraction. | 66<br><br>80% | O3AEE | Omacor® 1000mg/d<br>(EPA+DHA ~866mg/d)<br><br>Placebo (olive oil) | 46.8 | 1.24:1 |
| **Risk & Prevention (2013)**[15] | Italy | 12,505 | Patients w/multiple CV risk factors (at least 4 of the following: hypertension, hypercholesterolemia, current smoker, obesity, family history of premature CVD), clinical evidence of ASCVD, or any other condition placing the patient at high CV risk, but no MI. | 64<br><br>61% | EE | Omega-3 1000mg/d<br>(EPA+DHA ~870mg/d)<br><br>Placebo (olive oil) | 60 | 1:1.2 |
| **ASCEND (2018)**[16,17] | UK | 15,480 | Patients' w/T2DM but w/o ASCVD | 63.3<br><br>62% | EE | Omega-3 1000mg/d combined w/ 100mg/d aspirin (2x2)<br>(EPA 460mg/d + DHA 380mg/d)<br><br>Placebo (olive oil) | 88.8 | 1.2:1 |
| **GISSI-Prevenzione (2001)**[18] | Italy | 11,324 | Patients who had experienced a recent MI (within the previous 3 mos) and under-going preventive pharmacological interventions, alongside following Mediterranean dietary guidelines. | 59<br><br>85% | O3AEE | Omacor® 1000mg/d combined w/ 300mg/d vitamin E (2x2)<br>(EPA+DHA ~866mg/d)<br><br>Control (nothing given) | 42 | 1.24:1 |

| | | | | | | | | |
|---|---|---|---|---|---|---|---|---|
| **SU.FOL.OM3 (2010)[19]** | France | 2,501 | Patients' w/ a history of MI, unstable angina, or ischemic stroke in the previous year. | 61.4<br><br>79% | Unspecified | Omega-3 600mg/d combined w/ 560µg/d folate, 3mg/d vitaminB6, 20µg/d vitamin B12 (2x2)<br>(EPA 400mg/d + DHA 200mg/d)<br><br>Placebo (gelatin) | 56.4 | 2:1 |
| **ORIGIN (2012)[20]** | Multi-national | 12,536 | Patients' w/ T2DM, or impaired glucose tolerance, or prior MI, stroke, or revascularization, or angina w/ documented ischemia, or PAD, or microalbuminuria, or LVH. | 63.5<br><br>65% | O3AEE | Omacor® 1000mg/d combined w/ insulin glargine (2x2)<br>(EPA 465mg/d + DHA 375mg/d)<br><br>Control (standard care combined w/olive oil or insulin) | 74.4 | 1.24:1 |
| **Low Risk High Dose (LR-HD)** | | | | | | | | |
| **RCT-EPA (2023)[21]** | Canada | 121 | Men w/ prostate cancer who have has chosen to undergo radical prostatectomy. | 63<br><br>100% | MAG-EPA | Omega-3 3750mg/d<br>(EPA 3000mg/d)<br><br>Placebo (high oleic sunflower oil) | 14 | NA |
| **Mengelberg et al (2022)[22]** | New Zealand | 76 | Patients (60-90 yrs) w/ mild cognitive impairment. | 72.8<br><br>42% | TG | Omega-3 3000mg/d<br>(EPA 351mg/d +DHA 1491mg/d)<br><br>Placebo (linoleic acid) | 12 | 1:4.25 |
| **seAFOod Polyp Prevention (2019)[23]** | UK | 709 | Patients' (55-73 yrs) identified as 'high risk' of bowel cancer (detection of ≥5 small adenomas or ≥3 adenomas with at least one being ≥10 mm in diameter) after their 1st screening colonoscopy. | 65<br>80% | FFA/TG* | ALFA® x2 capsules/d (FFA) or 2780mg/d TGs combined w/ or w/o 300mg aspirin (2x2)<br>(EPA 2000mg/d)<br><br>Placebo (capric and caprylic acid medium-chain TGs) | 12 | NA |

| | | | | | | | | |
|---|---|---|---|---|---|---|---|---|
| **ENrGISE (2019)[24]** | USA | 289 | Patients aged ≥70 yrs w/ self-reported walking or stair-climbing difficulty and therefore at high risk for mobility disability and had evidence of low-grade chronic inflammation. | 78.3<br><br>52% | TG | Omega-3 1400-2800mg/d combined w/ 50-100mg/d losartan (2x2)<br>(EPA 800-1200mg/d + DHA 400-600mg/d)<br><br>Placebo (corn oil) | 12 | 2:1 |
| **ADCS-DHA (2010)[25]** | USA | 402 | Patients w/mild to moderate Alzheimer disease. | 76<br><br>47% | TG | Omega-3 2000mg/d<br>(DHA 2000mg/d)<br><br>Placebo (corn or soy oil) | 18 | NA |
| **MARINA (2011)†[26]** | UK | 163 | Non-smoking participants aged 45-70yrs w/ mild-to-moderate increased risk of CVD. | 55<br><br>39% | TG | Omega-3 1800mg/d<br>(EPA ~1083mg/d + DHA ~717mg/d)<br><br>Placebo (olive oil) | 24 | 1.51:1 |
| **Lin et al (2022)†[27]** | Taiwan | 312 | Patients ≥65yrs w/mild cognitive impairment or Alzheimer's Disease living in Veteran Retirement Centers. | 77.5<br><br>65% | EE | Omega-3 2000mg/d<br>(EPA 1600mg/d)<br><br>Placebo (isocaloric oil) | 24 | NA |
| **BRAVE-EPA (2025)[28]** | USA | 131 | Cognitively healthy male veterans aged 50-70 yrs at risk for Alzheimer's Disease. | 65.5<br><br>100% | IPE | Vascepa® 4000mg/d<br>(EPA 4000mg/d)<br><br>Placebo (unspecified) | 18 | NA |
| **EPOCH (2018)[29]** | New Zealand | 403 | Cognitively healthy community-dwelling adults aged 65-90 yrs. | 72.9<br><br>46% | rTG | Omega-3 2320mg/d<br>(EPA 600mg/d + DHA 1720mg/d)<br><br>Placebo (olive oil) | 18 | ~1:2.85 |
| **DREAM (2019)[30]** | USA | 535 | Patients w/ chronic, symptomatic moderate to severe dry eye disease. | 58<br><br>19% | TG | Omega-3 3000mg/d<br>(EPA 2000mg/d + DHA 1000mg/d)<br><br>Placebo (olive oil) | 12 | 2:1 |

| | | | | | | | | |
|---|---|---|---|---|---|---|---|---|
| **CAPFISH-3 (2024)**[31] | USA | 100 | Men on active surveillance for prostate cancer. | 64<br><br>100% | TG | Omega-3 2200mg/d combined w/ dietary counselling (low-fat diet) (2200mg/d EPA+DHA)<br><br>Control (nothing given) | 12 | Unspecified |
| **Low Risk Low Dose (LR-LD)** | | | | | | | | |
| **Shinto et al (2014)**[32] | USA | 67 | Patients aged ≥55yrs w/ probable Alzheimer dementia diagnosis. | 74.7<br><br>47% | TG | Omega-3 3000mg/d combined w/ 600mg lipoic acid (EPA 975mg/d + DHA 675mg/d)<br><br>Placebo (soybean oil) | 18 | 1.4:1 |
| **VITAL Rhythm (2021)**[‡33] | USA | 900 | Healthy adults aged ≥55yrs w/o history of CVD or cancer. | 66.7<br><br>50% | O3AEE | Omacor®1000mg/d combined w/ 2000IU/d vitamin D3 (2x2) (EPA 460mg/d + DHA 380mg/d)<br><br>Placebo (unspecified) | 63.6 | 1.24:1 |
| **CANN (2023)**[34] | UK, USA, and Australia | 259 | Older adults aged ≥55yrs w/ either subjective or mild cognitive impairment but without a diagnosis or evidence of dementia, or significant depression or other neurological disorders. | 65.5<br><br>43% | TG | Omega-3 1500mg/d combined w/ 500mg cocoa plant-derived flavanols (EPA 400mg/d + DHA 1100mg/d)<br><br>Placebo (corn and palm oil blend fatty acid composition typical of a UK or Australian diet) | 12 | 1:2.8 |
| **AREDS2 (2013)**[35] | USA | 4,203 | Patients aged 50-85 yrs w/intermediate advanced age-related macular degeneration (AMD) in both eyes or intermediate AMD in one and advanced AMD in the other eye. | 74<br><br>43% | AREDS2 | AREDS2 2x capsules/d (EPA 650mg/d + DHA 350mg/d combined w/ 500 mg/d vitamin C, 440IU/d vitamin E, 15mg/d beta-carotene, 80mg/d zinc oxide and 2mg/d cupric oxide (2x2).<br><br>Control: standard AREDS supplement w/o omega-3. | 57.6 | ~1.9:1 |

| | | | | | | | | |
|---|---|---|---|---|---|---|---|---|
| **MARINA (2011)**[†26] | UK | 163 | Non-smoking participants aged 45-70yrs w/ mild-to-moderate increased risk of CVD. | 55<br><br>39% | TG | Omega-3 450mg/d<br>(EPA ~270mg/d + DHA ~179mg/d) or<br>Omega-3 900mg/d<br>(EPA ~542mg/d + DHA ~359mg/d)<br><br>Placebo (olive oil) | 24 | 1.51:1 |
| **Lin et al (2022)**[†27] | Taiwan | 312 | Patients ≥65yrs w/mild cognitive impairment or Alzheimer's Disease living in Veteran Retirement Centers. | 77.5<br><br>65% | EE | Omega-3 1000mg/d<br>(DHA 700mg/d) or<br>Omega-3 2000mg/d<br>(EPA+DHA 1150mg/d)<br><br>Placebo (isocaloric oil) | 24 | Unspecified. |
| **DO-HEALTH (2025)**[36] | Europe | 2157 | Community-dwelling adults aged ≥70 yrs generally healthy and active, w/no major health events (e.g., cancer or MI) in the 5 yrs prior to enrollment. | 74<br><br>38% | TG | Omega-3 1000mg/d combined w/ 2000IU/d vitamin D3 and strength exercise 3x /wk. (2x2)<br>(EPA 330mg/d + DHA 660mg/d)<br><br>Placebo (sunflower oil) | 36 | 1:2 |

Abbreviations: AREDS2: Age-Related Eye Disease Study 2 formulation; ASCVD: atherosclerotic cardiovascular disease; CAD: coronary artery disease; CVD: cardiovascular disease; DHA: docosahexaenoic acid; EE: ethyl ester; EPA: eicosapentaenoic acid; FFA: free fatty acid; HDL: high-density lipoprotein; HR-HD: high risk, high dose; HR-LD: high risk, low dose; IPE: icosapent ethyl; LR-HD: low risk, high dose; LR-LD: low risk, low dose; LVH: left ventricular hypertrophy; MAG: monoacylglyceride; mg/d: milligram per day; MI: myocardial infarction; mos: months; N: number of participants randomized; NA: not applicable; O3AEE: omega-3 acid ethyl esters; O3CA: omega-3 carboxylic acids; PAD: peripheral artery disease; RCT: randomized controlled trial; rTG: re-esterified triglyceride; TD1/2: type 1/2 diabetes mellitus; TG: triglyceride; UK: the United Kingdom; USA: the United States of America; wks: weeks; yrs: years

*In seAFOod Polyp Prevention trial, the omega-3 intervention formulation was modified mid-trial due to discontinuation of the original product. Participants initially received 2000 mg/day of >99% pure EPA (FFA); when supply ceased, ~one-third were switched to an equivalent EPA dose provided as 2780 mg/day of 90% EPA in TG form.

†MARINA and Lin et al., investigated multiple doses of omega-3 in low-risk participants. As a result, their low-dose arms were included in the LR-LD category, and their high-dose arms were included in the LR-HD category in the stratified analyses.

‡VITAL-rhythm is an ancillary trial of the VITAL trial (NCT01169259)- our meta-analysis was performed using VITAL-rhythm data.
