## Supplementary Appendix 1 for "Effects of Omega-3 Fatty Acid Treatment on Risk for Atrial Fibrillation: An Updated Meta-Analysis of 34 Trials including 114,326 Individuals"

**Supplementary Appendix 2.** Rationale for setting an intake of 1500 mg/d for EPA/DHA to differentiate dietary/supplement intake levels from pharmaceutical dose levels

**Ecological data**. The age-standardized global prevalence of AF in 2021 was reported in a 2025 update from the American Heart Association (1). The rate of AF in the US, which has an average Omega-3 Index (O3I, erythrocyte EPA+DHA%) of 5% (2), is >1000 pr 100,000 people. Compared with this, Japan, a country with an average O3I of ~10% (2), has an AF rate of <400 per 100,000. In a 2000 publication, Sugano stated, “Fish and shellfish now provide 5–6 g fat and **<1–2 g** n3 PUFA per person per day in Japan.” (3) Clearly, there are multiple differences between the US and the Japanese populations, but this observation is at least consistent with the hypothesis that higher intakes and tissue levels of omega-3 PUFAs are associated with lower risk for AF. Importantly, data from a prospective cohort study in Japan including about 42,000 individuals between 40 and 59 years of age reported that the median EPA+DHA intake in the cohort was **900** mg/d, and the intake in the highest quintile – where risks for total MI and nonfatal coronary events were the lowest - was **2100** mg/d (4). AF risk was not tracked in this report.

**Omega-3 Intake data**. A large observational study from Denmark found that consumption of around **600** mg/d of EPA+DPA+DHA (quintile 3, Q3; **Table 1**) was associated with the lowest risk of AF compared with lower and higher levels, i.e., there was a U-shaped relationship (5). However, intakes in Q4 and Q5 were not significantly different from Q1 (reference) in any of the three models tested (**Table 1**). Q5 was >**0.99** g/d. Data from the Million Veterans study (6) suggested that risk for AF was reduced maximally at about **500 mg/d** but remained significantly reduced up to **~1300** mg/d of EPA+DHA (**Figure 1**).

**Table 1. Associations between quintiles of omega-3 intake and risk for AF. Taken from Rix et al.**

| | Crude | | | Primary adjusted model[a] | | | Secondary model[b] | | |
|---|---|---|---|---|---|---|---|---|---|
| | HR | 95% CI | P | HR | 95% CI | P | HR | 95% CI | P |
| Q1 <0.39 g/day | 1 | – | – | 1 | – | – | 1 | – | – |
| Q2 0.39–0.53 g/day | 0.92 | 0.82–1.03 | 0.14 | 0.92 | 0.82–1.03 | 0.16 | 0.92 | 0.82–1.04 | 0.18 |
| Q3 0.54–0.73 g/day | 0.87 | 0.77–0.97 | 0.02 | 0.87 | 0.78–0.98 | 0.02 | 0.87 | 0.78–0.98 | 0.02 |
| Q4 0.74–0.99 g/day | 0.95 | 0.85–1.07 | 0.40 | 0.96 | 0.86–1.08 | 0.49 | 0.96 | 0.86–1.08 | 0.52 |
| Q5 >0.99 g/day | 1.06 | 0.95–1.18 | 0.30 | 1.05 | 0.93–1.18 | 0.42 | 1.05 | 0.93–1.18 | 0.43 |

PUFA, polyunsaturated fatty acids; HR, hazard ratio; CI, confidence interval; Q, quintile.
[a]Adjusted for age, sex, hypertension, systolic blood pressure, body-mass index, waist circumference, smoking, alcohol intake, years in school, hypercholesterolaemia and/or cholesterol treatment, total serum cholesterol, angina pectoris, diabetes mellitus, myocardial infarction, heart failure, and total energy intake.
[b]Sensitivity analysis further adjusted for consumption of fruits, vegetables, red meat, poultry, and fatty dairy products.

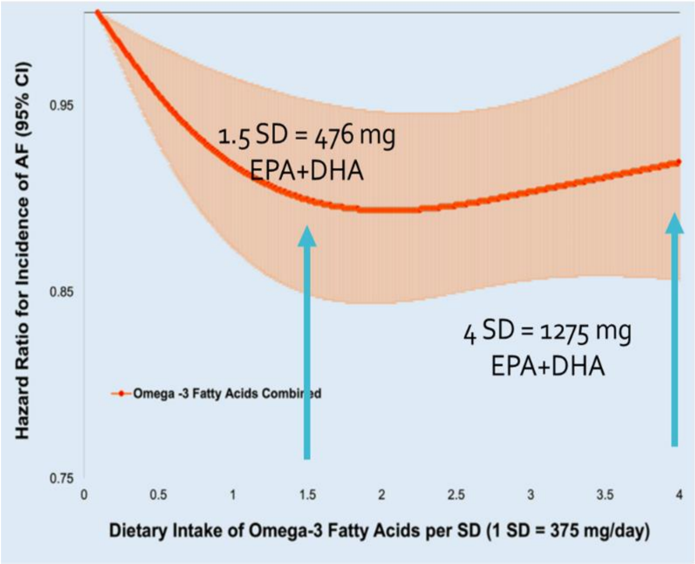


Figure 1. Association between reported EPA+DHA intake and risk for AF in a Veterans cohort. Risk for AF was significantly and maximally reduced (compared with no intake) between about 476 and 1270 mg EPA+DHA per day. From Guardino et al.

**Fish oil supplement use data**. An alternative approach to this question is to determine the extent to which reported fish oil supplement (FOS[1]) use is associated with risk for AF. Two such studies (from the same research group) have been reported in the UK Biobank (UKBB). Both Zhang et al. (7) and Chen et al. (8) concluded that the use of FOS was associated with an increased risk (10-13%) for AF. (Importantly, Chen et al. also reported that FOS use was favorably associated with five other CVD outcomes in the same study). These findings have been re-examined by O'Keefe et al. (9) and found to be in error, that being the failure to adjust for age properly (i.e., continuously, instead of dichotomously as was done in the earlier studies). Thus, there is no association between FOS use and risk for AF in the UKBB.

**Blood omega-3 data.** As dietary intake (and FOS use) data are subject to a number of uncertainties (faulty memory, old FA data in food databases, compliance with and doses of FOS, etc.), biomarkers of omega-3 PUFA status (whether in plasma, plasma phospholipids, whole blood and/or erythrocytes) provides a more objective measure of tissue exposure. Accordingly, two recent studies from the Fatty Acids and Outcomes Research Consortium (FORCE group) are important in this regard. First, a meta-analysis of de novo data from 17 observational cohorts around the world including about 55,000 individuals found that those individuals with the highest EPA+DHA levels (compared with the lowest; 90th vs 10th percentiles), were 12% less likely to develop AF after a median follow-up of 13 years (**Figure 2**) (10). The second report took the same approach but focused on risk for stroke. That study included data from 29 cohorts and over 183,00 individuals followed for a median of 14 years. As was seen with AF, the risk for stroke was significantly reduced in a dose dependent manner

[1] Any effects of or relationships with fish oil supplement use could reasonably be extended to the use of any dietary supplement providing EPA and/or DHA such as algal oil, krill oil, cannalus oil, green lipped muscle oil, etc.).

(**Figure 3**)(11). In both of these studies, the estimated O3I of the highest quintile was about 8% compared to <4% in the reference group.

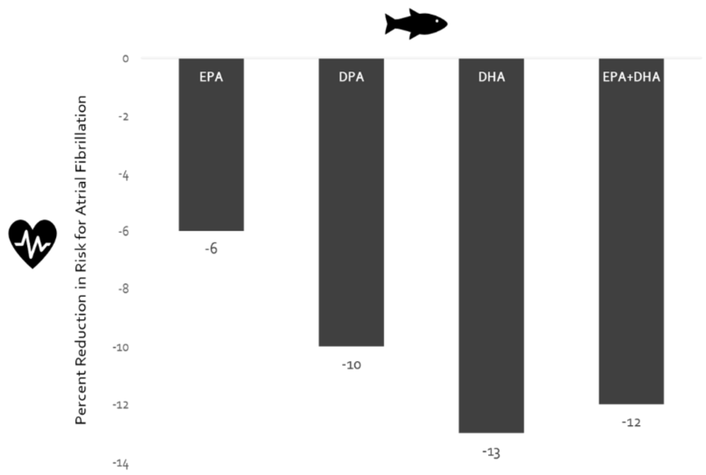


Figure 2. Association between circulating n-3 PUFA levels and odds for atrial fibrillation. From Qian et al.

Second, a study conducted only within the UK Biobank (UKBB) confirmed the findings of Qian et al. That study (9) included data on 261,108 UKBB participants followed for 12.7 years for new onset AF. The higher levels (quintiles 2-5) for total omega-3, DHA and non-DHA omega-3 were all significantly reduced compared with quintile 1 (**Figure 4**). When these new data were combined with the original meta-analysis of Qian et al., the relationships between omega-3 levels and risk for AF were confirmed and strengthened (**Table 2**). Multiple other FORCE studies comparing disease outcomes as a function of blood omega-3 levels have consistently shown the greatest protection at the highest blood levels (**Figure 5**), again levels associated with intakes of up to 1500 mg/d.

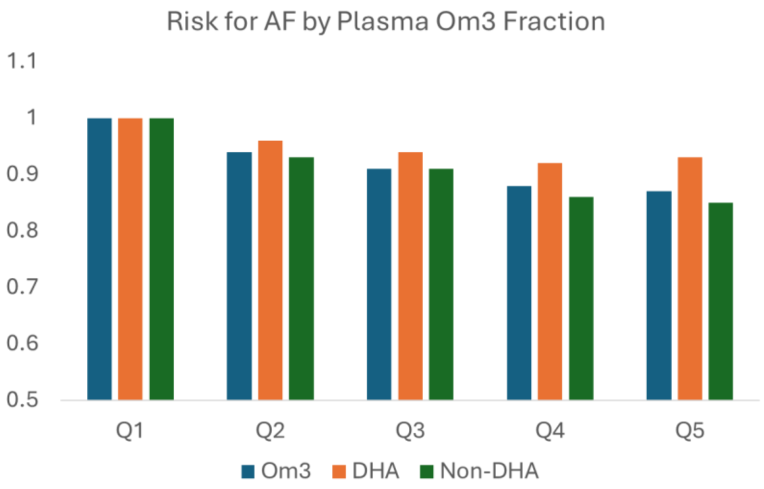


Figure 4. Association between plasma n-3 PUFA levels and odds for atrial fibrillation. All Q2-Q5 HRs lower than Q1 (p<0.01). From O'Keefe (ms under review JAHA 7/2025).

**Randomized trials with fish oils in the general population.** The only large RCT to address the question of how omega-3 supplementation affects risk for AF is the VITAL study (12). Here 25,119 men and women over age 50 were randomized to **840** mg EPA+DHA/day (and/or vitamin D) and followed for incident AF for 5.3 years. The risk for AF was 9% higher in the treated group than the placebo group but this difference was not statistically significant (p=0.19). Thus, there was no reduction in risk for AF as the longer-term observational data suggested, but at least in this group of older patients treated for 5 years with omega-3, there was no increase in risk for AF. Hence, the safety of that dose vis-à-vis AF was confirmed as also suggested by the FOS use studies in the UKBB.

Table 2. Association between plasma n-3 PUFA levels and odds for atrial fibrillation. All Q2-Q5 HRs lower than Q1 (p<0.01)

| | Analysis | Adjusted Hazard Ratio[1] |
|---|---|---|
| DHA | Original meta-analysis | 0.90 (0.85, 0.96) ** |
| | UK Biobank alone | 0.93 (0.89, 0.97) *** |
| | **Updated meta-analysis** | **0.92 (0.89, 0.96) *** ** |
| EPA | Original meta-analysis | 1.00 (0.95, 1.05) |
| | UK Biobank alone | 0.92 (0.88, 0.95) *** |
| | **Updated meta-analysis** | **0.94 (0.92, 0.97) *** ** |
| DPA | Original meta-analysis | 0.89 (0.83, 0.95) *** |
| | UK Biobank alone | 0.91 (0.87, 0.94) *** |
| | **Updated meta-analysis** | **0.90 (0.88, 0.93) *** ** |
| EPA + DHA | Original meta-analysis | 0.93 (0.87, 0.99) * |
| | UK Biobank alone | 0.92 (0.88, 0.95) *** |
| | **Updated meta-analysis** | **0.92 (0.90, 0.95) *** ** |

*Further considerations of the EPA+DHA intake that does not increase the risk for AF.*

However, estimates can also be made based on the blood biomarker data from the two FORCE studies, Qian et al. (10) and O'Keefe et al. (9). In the former, the authors noted that the mean (SD) marine omega-3 intake in the 10 countries from which the 17 cohorts were drawn was about **430 (350)** mg/day (13). Assuming a normal distribution (which it probably is not), the 95$^{th}$ percentile intake (mean + 2 SD) would be **1130** mg/day. In the latter study, the estimated O3I in the highest EPA+DHA quintile was about 8% [which is consistent with the originally-proposed omega-3 index target of >8% for minimizing risk of CVD death (14)]. In the UKBB, where the highest quintiles of plasma omega-3 were associated with the lowest risk of both AF and stroke, the estimated O3I was 7-8%. Thus, what intake is associated with an O3I of around 8%? Unfortunately, the actual intakes of omega-3 across the quintiles of plasma omega-3 in the UKBB are not known. However, we know that a dose of **1800** mg/day of EPA+DHA was shown to increase the O3I from 4.3% to 9.5% after 5 months of treatment, and a dose of **900** mg/day raised the O3I to 7.5% (15).

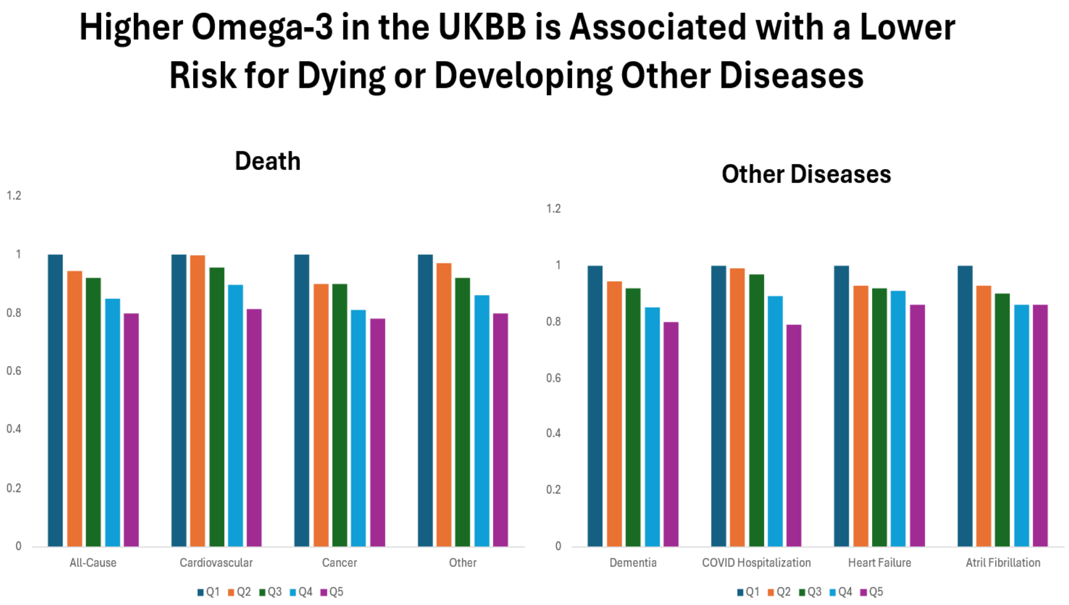


Figure 5. Summary of relationships between quintiles of blood omega-3 levels and several incident diseases from the UKBB

In 2021, the typical adult American ate ~1.5 serving/week of fish or seafood (16); and the average intake of DHA + EPA in the United States is approximately 100 mg/d (17, 18). This corresponds to an average DHA + EPA level in red blood cells (i.e., the omega-3 index) in the United States of about 5.4% (19). The estimated median omega-3 index in Q5 in the Qian study was 8.62%. A pooled analysis of individual-level data from 14 omega-3 supplementation studies allowed for the construction of an equation that would predict the amount of EPA+DHA needed to achieve an O3I of 8% from different starting levels (18). Using this equation, the amount of EPA+DHA needed to raise an *average* O3I of 5.4% to 8% is **900** mg/d. For 95% of the population (not just 50%) to achieve an O3I of 8% would require an increase of approximately **1500** mg/d of DHA + EPA (17, 20).

In light of these considerations and recognizing that the lowest approved dose of an omega-3 pharmaceutical product (Epadil) is **1800 mg/day**, we consider that a reasonable dividing line between "low" intakes from food and/or dietary supplements and "high" doses used in drug studies would be ~1500 mg EPA+DHA per day. This value was thus applied to the updated meta-analysis of omega-3 RCTs and risk for AF. As noted above, an intake of ~1500 mg/d is also associated with the highest quintile omega-3 index levels, and these levels are linked with the lowest risk for AF [not to mention stroke (11) and total mortality (21)].
