## Supplementary Appendix 2 for "Effects of Omega-3 Fatty Acid Treatment on Risk for Atrial Fibrillation: An Updated Meta-Analysis of 34 Trials including 114,326 Individuals"

**Supplementary Appendix 2.** Funding and acknowledgement statements for RCTs included in meta-analysis

'AZD0585 Phase III Long-term Study in Japan' (NCT02463071) by Yokote et al (2020) was funded by AstraZeneca K.K.

REDUCE- IT (NCT014923615) trial and analysis funded by Amarin.

STRENGTH (NCT02104817) trial was funded by AstraZeneca AB and coordinated by the Cleveland Clinic Coordinating Center for Clinical Research (C5Research).

FAAT (NCT00004559) study was supported in part by a grant from the NHLBI, NIH (HL62154). Other contributors to FAAT are Jeremy Ruskin, Massachusetts General Hospital, Boston; David Martin, Lahey Clinic, Burlington, Mass; Alfred E. Buxton, Rhode Island Hospital, Providence; Peter Holzberger and Mark L. Greenberg, Dartmouth-Hitchcock Medical Center, Lebanon, NH; Bruce Stambler, University Hospitals of Cleveland, Cleveland, Ohio; Jonathan S. Steinberg, St. Lukes–Roosevelt Hospital, New York, NY; Brian Olshansky, University of Iowa Hospitals, Iowa City; and Bruce Lerman, New York Hospital–Cornell Medical Center, New York, NY.

PEACH (UMIN000003171) study was funded by the Japan Heart Foundation (No. 12090021).

OMEMI (NCT01841944) study was supported by grants from Stein Erik Hagen Foundation to Center for Clinical Heart Research, Oslo, Norway; Olav Thons Foundation, Oslo, Norway and Tom Wilhelmsen Foundation, Oslo, Norway. The grants were unrestricted and the Foundations did not have any impact on any part of the study, i.e. data collection, analysis, and interpretation of data or in writing the manuscript. Study medication and placebo were sponsored by Orkla Health, Oslo Norway, which also had no impact on any part of the study.

EVAPORATE trial (NCT02926027) was funded by Amarin Pharma, Inc. (Bridgewater NJ, USA).

The HEARTS trial (NCT01624727) was supported by the National Institutes of Health Specialized Center of Clinically Oriented Research program grant to Dr Welty (P50

HL083813) and by the Harvard Clinical and Translational Science Center Award (NIH UL1 TR001102).

No sources of funding or external support were disclosed in the SAREPITASO (SARpogrelate-EPA-PITavastatin-ASO) study.

Derosa et al. (2016) reported no funding, sponsorship, or support acknowledgements in their study examining the effects of n-3 PUFAs on fasting plasma glucose and insulin resistance in individuals with IFG or IGT."

OFAMI (NCT01422317) supported by Pharmacia-Upjohn A/S, Oslo, and by Pronova A/S, Oslo.

RESPECT EPA (UMIN000012069) study was supported by a grant from the Japan Heart Foundation, Japan.

GISSI-HF (NCT00336336) trial was supported by grants from Societa´ Prodotti Antibiotici (SPA; Italy), Pfizer, Sigma Tau, and AstraZeneca.

Risk and Prevention Study (NCT00317707) supported by Società Prodotti Antibiotici, Pfizer, and Sigma-Tau.

ASCEND study (NCT00135226) supported by grants to the University of Oxford from the British Heart Foundation. The Clinical Trial Service Unit at the University of Oxford receives support from the U.K. Medical Research Council (which funds the Medical Research Council Population Health Research Unit in a strategic partnership with the University of Oxford), the British Heart Foundation, and Cancer Research U.K. Bayer (Germany) provided aspirin and placebo tablets that were used in the trial, and Solvay, Abbott, and Mylan provided the n−3 fatty acids and placebo. The companies and Bayer U.S. provided some funding for drug packaging.

GISSI-Prevenzione was supported by grants from Bristol-Myers Squibb and Pharmacia.

The SU.FOL.OM3 study (ISRCTN41926726) acknowledges financial support from the French Ministry of Research (R02010JJ), Ministry of Health (DGS), Sodexo, Candia, Unilever, Danone, Roche Laboratory, Merck EPROVA GS and Pierre Fabre Laboratory.

ORIGIN trial (NCT00069784) supported by Sanofi, with study drugs provided by Pronova BioPharma Norge.

RCT-EPA (NCT02333435) was mainly supported by the Canadian Cancer Society (Grant #702569). It was also supported by the Foundation of CHU de Québec-Université Laval. RCT EPA investigators also thank Samuel Fortin from SCF Pharma (Ste-Luce, Qc) for having kindly provided the MAG-EPA and placebo HOSO capsules. The authors thank Line Berthiaume for fatty acid profiling.

'In older adults with mild cognitive impairment, do omega-3 fatty acids compared to a placebo improve cognitive performance?' study (ACTRN12614000874617) by Mengelberg et al (2022) was supported by funding from the Oakley Mental Health Research Foundation, the Neurological Foundation, HOPE Selwyn Foundation scholarships for Research on Ageing, and a Massey University Research Fund.

The seAFOod Polyp Prevention trial (ISRCTN05926847) was funded by the Efficacy and Mechanism Evaluation Programme, a partnership between the UK Medical Research Council and the National Institute for Health Research (NIHR; project 09/100/25). The views expressed are those of the author(s) and not necessarily those of the NIHR or the Department of Health and Social Care.

The ENRGISE Study (NCT02676466) was funded by a National Institutes of Health/ National Institute on Aging Cooperative Agreement U01AG050499. The research is partially supported by the Claude D. Pepper Older Americans Independence Centers at the University of Florida (P30AG028740), Wake Forest University (P30AG021332), Tufts University (P30AG031679), and University of Pittsburgh (P30AG024827). Tufts University is also supported by the Boston Rehabilitation Outcomes Center (R24HD065688-01A1). Abbott Laboratories provided funding for the purchase of study drug and matching placebo.

ADCS-DHA (NCT00440050) study was supported by grant UO1-AG10483 from the National Institute on Aging. The placebo and DHA study drugs were provided by Martek Biosciences. Martek also provided plasma and cerebrospinal fluid measurements of fatty acids, as well as partial financial support for the magnetic resonance imaging sub study.

The MARINA trial (ISRCTN66664610) funding was provided by the Food Standards Agency and the Department of Health, England (Project code N02041).

'The Protective Effect of Omega-3 Fatty Acid on Cognitive Function Among Patients With Mild Dementia' study (NCT04972643) by Lin et al (2022) were supported by the following grants: MOST 109-2320-B-038-057-MY3, 109-2320-B-039-066, 110-2321-B-006-004, 110-2811-B-039-507, 110-2320-B-039-048-MY2, and 110-2320-B-039-047-MY3 from the Ministry of Science and Technology, Taiwan; NHRI-EX108-10528NI from the National Health Research Institutes, Taiwan; MYRG2018-00242-ICMS from University of Macau, China; ANHRF109-31, 110-13, and 110-26 from An Nan Hospital, China Medical University, Tainan, Taiwan; CMRC-CMA-3 from Higher Education Sprout Project by the Ministry of Education (MOE), Taiwan; CMU108-SR-106 from the China Medical University, Taichung, Taiwan; and CMU104-S-16-01, CMU103-BC-4-1, CRS-108-048, DMR-108-216, DMR-109-102, DMR-109-244, DMR-HHC-109-11, DMR-HCC-109-12, DMR-HHC-110-10, DMR-110-124, and CMU110-AWARD-02 from the China Medical University Hospital, Taichung, Taiwan.

BRAVE-EPA (NCT02719327) study funding was provided by VA Office of Research and Development (CX001261). Infrastructure provided by the Wisconsin Alzheimer's Disease Research Center (ADRC) was supported through NIA P30 AG062715. Icosapent ethyl was provided by Amarin Pharma, Inc. (Bridgewater, NJ, USA). Neither the funding agencies nor Amarin had input in analysis, endpoint adjudication, study performance or measures. The contents do not represent the view of the VA or the United States Government.

The EPOCH trial (ACTRN2607000278437) was funded by the National Health and Medical Research Council of Australia and CSIRO.

DREAM (NCT02128763) study supported by cooperative agreements U10EY022879 and U10EY022881 from the National Eye Institute, National Institutes of Health, Department of Health and Human Services. Additional support provided by the Office of Dietary Supplements National Institutes of Health, Department of Health and Human Services. The sponsor participated in overseeing the conduct of the study. EPAX Norway AS (Aalesund, Norway) contributed the fish oil and the Access Business Group, LLC (Ada, MI) manufactured, packaged, and delivered study supplements to the central pharmacy. OCULUS Inc. (Arlington, WA) provided discounted equipment, customized software, and user support for the keratography used in the study. RPS Diagnostics, Inc. (Sarasota, FL) provided InflammaDry Detector test kits to the clinical centers for testing MMP-9. TearLab Corporation (San Diego, CA) provided a discount to the study for testing materials for their TearLab Osmolarity System.

The CAPFISH-3 trial (NCT02176902) is supported by a grant from National Cancer Institute at the National Institute of Health (RO1CA231219).

'Lipoic Acid and Omega-3 Fatty Acids for Alzheimer's Disease' study (NCT01058941) by Shinto et al (2014) was supported by the National Institutes of Health/National Institute of Aging (NIH/NIA) R21AG023805, NIH/NIA AG08017, and NIH General Clinical Research Grant M01RR00334. Nordic Natural, Watsonville, CA, USA, supplied the fish oil and placebo oil, and Meda Pharma, Bad Homburg, Germany, supplied the lipoic acid and placebo.

The VITAL Rhythm Study (NCT01169259) was supported by grant R01HL116690 from the National Heart, Lung, and Blood Institute and the VITAL trial was supported by grants U01 CA138962 and R01 CA138962, which included support from the National Cancer Institute, National Heart, Lung and Blood Institute, Office of Dietary Supplements, National Institute of Neurological Disorders and Stroke, and National Center for Complementary and Integrative Health. Pharmavite LLC (vitamin D3) and Pronova BioPharma/BASF (omega-3 fatty acids) donated the study agents, matching placebos, and packaging.

The CANN Study (NCT02525198) was funded in part by Abbott Nutrition via a Centre for Nutrition, Learning, and Memory (CNLM) grant to the University of Illinois, which in turn awarded a research grant to the authors through a competitive peer reviewed process. Study foods were provided free of charge by Abbott Nutrition (Columbus, OH, United States).

The AREDS2 trial (NCT00345176) was supported by the intramural program funds and contracts from the National Eye Institute/National Institutes of Health (NEI/NIH), Department of Health and Human Services, Bethesda, MD, Contract No. HHS-N-260-2005-00007-C and ADB Contract No. N01-EY-5-0007. Funds were generously contributed to these contracts by the following NIH institutes: Office of Dietary Supplements (ODS), National Center for Complementary and Alternative Medicine

(NCCAM), National Institute on Aging (NIA), National Heart, Lung and Blood Institute (NHLBI), and National Institute of Neurological Disorders and Stroke (NINDS).

The DO-HEALTH study (NCT01745263) was funded by the Seventh Framework Program of the European Commission (grant agreement 278588; principal investigator Heike A. Bischoff-Ferrari, University of Basel, Switzerland) and several industry partners within the EU Framework including DSM Nutritional Products, Roche, NESTEC, Pfizer, and Streuli.
